# Dynamic shift in the dominant transmission route of clade Ib mpox virus across networks with sexual and non-sexual contacts

**DOI:** 10.1101/2025.07.18.25331780

**Authors:** Fuminari Miura, Ka Yin Leung, Maria Xiridou, Marten van Antwerpen, Nicola Low, Niel Hens, Emmanuel Hasivirwe Vakaniaki, Jacco Wallinga

## Abstract

The intensifying outbreaks of the novel monkeypox virus clade Ib in the Democratic Republic of the Congo have raised global concern about the potential for wider epidemic spread. Some clade Ib mpox outbreaks have shown a distinct transmission pattern in which transmission associated with both sexual and non-sexual contacts co-exist. Here, we characterize these outbreaks in a network epidemic model, which incorporates sexual and non-sexual contacts, and project age- and route-specific transmission potentials under a wide range of scenarios. Our analyses suggest that the dominant route of transmission may shift over time from sexual to non-sexual contacts, which leads to larger epidemics. The age groups contributing most to overall infections and mortality also change over time, suggesting that target groups for intervention should be adjusted accordingly. For countries at risk of travel-associated mpox outbreaks, these findings highlight the importance of monitoring evolving monkeypox virus transmission patterns and interacting transmission routes to support timely and effective control measures.

**Significance statement:** The initial outbreak of clade Ib monkeypox virus in the Democratic Republic of Congo was characterized by differences in how the virus spreads, with transmission through both sexual and non-sexual contacts playing a crucial role. This study addresses two key public health challenges: how does an outbreak evolve when multiple transmission routes coexist, and how to translate that understanding into effective interventions. We develop a mathematical framework to address these challenges and show that the dominant transmission route can shift over time, from sexual to non-sexual. We show that even small increases in transmission through non-sexual contacts can lead to substantially larger epidemics, underscoring the need for continuous monitoring and adaptive response strategies.

**One sentence summary:** During an mpox outbreak, the dominant route of transmission and optimal target groups for intervention may shift over time

## Introduction

The emergence of clade Ib monkeypox virus (MPXV) in the Democratic Republic of the Congo (DRC) in late 2023 has raised renewed global concern about the potential for wider epidemic spread (1). In August 2024, the World Health Organization declared a second Public Health Emergency of International Concern about mpox in response to escalating outbreaks in Central and East Africa, driven by this novel subclade (2). Initially identified in Kamituga health zone, South Kivu province, clade Ib has since spread to multiple neighboring countries and travel-associated cases have been reported in Europe, the Americas, and Asia (3). Although sustained transmission of clade Ib outside Africa has not been confirmed, as of June 2025, the combination of uncertain epidemiology, likely under-detection, and limited control capacity in affected regions make further national and international spread possible (4). Several risk assessments are now underway in Europe and other regions to evaluate the potential impact of future introductions and to guide preparedness planning (5, 6).

The transmission dynamics of clade I MPXV are changing. MPXV is historically a zoonotic pathogen, with clade Ia circulating through animal-to-human and subsequent limited human-to-human transmission (7). In contrast, the outbreaks of clade Ib have been characterized by sustained human-to-human transmission via close contact either within households and communities (“non-sexual” route) or in association with sexual encounters (“sexual” route). Studies based on data collected in South Kivu in 2024-2025 suggest that key epidemiological parameters for clade Ib MPXV vary by transmission route; for instance, incubation periods and serial intervals may be shorter for transmission via sexual contacts than via non-sexual contacts (8, 9). However, secondary attack risks (SARs), reproduction numbers, and other key parameters remain poorly quantified, especially in different age groups. Case fatality ratios (CFRs) for clade I MPXV have historically ranged from 1– 10% (7, 10), with recent crude estimates for clade Ib ranging from 0.2–0.5% (3). These CFRs appear to be higher in young children and lower in adults, although differences across age and clade may reflect variation in surveillance capacity or access to clinical care.

Capturing the dynamics of mpox within and across networks of sexual and non-sexual contacts may help to design efficient control strategies. Clinical and epidemiological investigations suggest that the initial phase of the clade Ib outbreak in Kamituga was driven by transmission via sexual contacts between women and men (1, 11), with subsequent transmission from person-to-person via close physical contacts in the household or community among children under 15 year old (12). Following spread of clade Ib MPXV to Burundi, the age and sex distribution of mpox cases there also suggests spread through both sexual and non-sexual contacts (13). This pattern contrasts with the 2022 global outbreak of clade IIb mpox, where transmission via sexual contacts occurred mainly between men who have sex with men and transmission within the household to younger age groups was minimal (14, 15). In Sierra Leone, however, where a large clade IIb MPXV outbreak started in 2025, the distribution of cases also suggests transmission via both sexual and non-sexual contacts (16).

Multiple transmission routes, including through sexual contact, have been described for emerging infectious diseases, such as Ebola (17) and Zika (18). Existing mathematical models describing transmission via both sexual and non-sexual contacts often simplify transmission via sexual contacts by ignoring network structure and assuming individuals only contact each other in one-off encounters for Zika and Ebola (19–21) and mpox (22). Network models that explicitly capture sexual contact networks are more realistic (23–25) but their application in outbreak responses remains limited. This is largely due to their added complexity and challenges in linking model structure to epidemiological data collected during outbreaks (26, 27).

We developed a network epidemic model tailored to clade Ib mpox, using the Kamituga outbreak as a motivating example, where multiple, age-structured contact routes of human-to-human transmission have interacted and evolved over time. Building on prior frameworks (23, 24, 28), our model incorporates both sexual and non-sexual contact structures; non-sexual contacts are defined as age-specific contacts and are parameterized by a contact matrix (29). Sexual contacts are modeled to occur within pairs of sexually active individuals (partnerships), forming a static sexual network according to the distribution of the number of sexual partners.

The primary objective of this study is to determine how the dominant route of human-to-human transmission evolves over time, using epidemiological data that can be collected during outbreaks of clade Ib MPXV. We project age- and route-specific contributions to transmission at different epidemic stages as well as epidemic sizes by transmission route. We also show how the model can be used to identify which subgroups would be most effective to target with interventions, based on their projected contributions to onward transmission and mortality (29–31). Finally, although motivated by ongoing clade Ib outbreaks, the proposed framework aims to be broadly applicable to infections transmitted through both sexual and non-sexual contacts. In particular, we explore a wide range of scenarios to reflect differential epidemiological and behavioral conditions across mpox outbreak settings, not necessarily limited to clade Ib. This makes our study generic and relevant for supporting risk assessment not only in currently affected countries such as the DRC, but also in countries at risk of future mpox importation, including those outside the African region.

## Results

### Projected transmission potential of each transmission route

To project route- and age-specific transmission potentials, we used a network epidemic model with two routes of transmission via close contact – sexual and non-sexual – including age differences while simplifying by ignoring any sex differences. The two transmission routes differed in the contact patterns and the route-specific SARs. We computed a projection matrix that has the number of new infections produced by each age group and route of transmission as its elements (i.e., the next-generation matrix (NGM) (29, 32)). The NGM can be subdivided into four submatrices, which describe transmission from one route to another (via either sexual or non-sexual contact, indexed as *s* and *ns* respectively). Each submatrix can be summarized as a route-to-route specific reproduction number; *R*_*s,ns*_ represents the expected number of new infections via sexual contacts transmitted by a case who acquired infection via non-sexual contact (‘from-non-sexual-to-sexual’ reproduction number) in the absence of other transmission routes. Similarly, *R*_*s,s*_, *R*_*ns,s*_, and *R*_*ns,ns*_ correspond to reproduction numbers for ‘from-sexual-to-sexual’, ‘from-sexual-to-non-sexual’, and ‘from-non-sexual-to-non-sexual’, respectively.

The submatrix of the NGM for transmission from-non-sexual-to-non-sexual contacts projects higher secondary infections among young children or from children to older adults in their 20s and 30s (**Figure 1(A)**), while secondary infections via transmission from-sexual-to-sexual contacts were mainly produced by younger adults aged 15-19 (**Figure 1(B)**), using a baseline scenario with parameterization of the model for Kamituga (see the details in **Methods** and **Supplementary Materials**). We can quantify the relative contribution of transmissions within and between the two routes by projecting reproduction numbers for each submatrix under the baseline scenario (**Figure 1(C)**). To determine the overall transmission potential *R*_0_ (the basic reproduction number, where *R*_0_ > 1 indicates that an outbreak will grow), the full NGM is required to capture the interaction within and between transmission routes (see the proof in **Supplementary Materials**).

**Figure 1.**
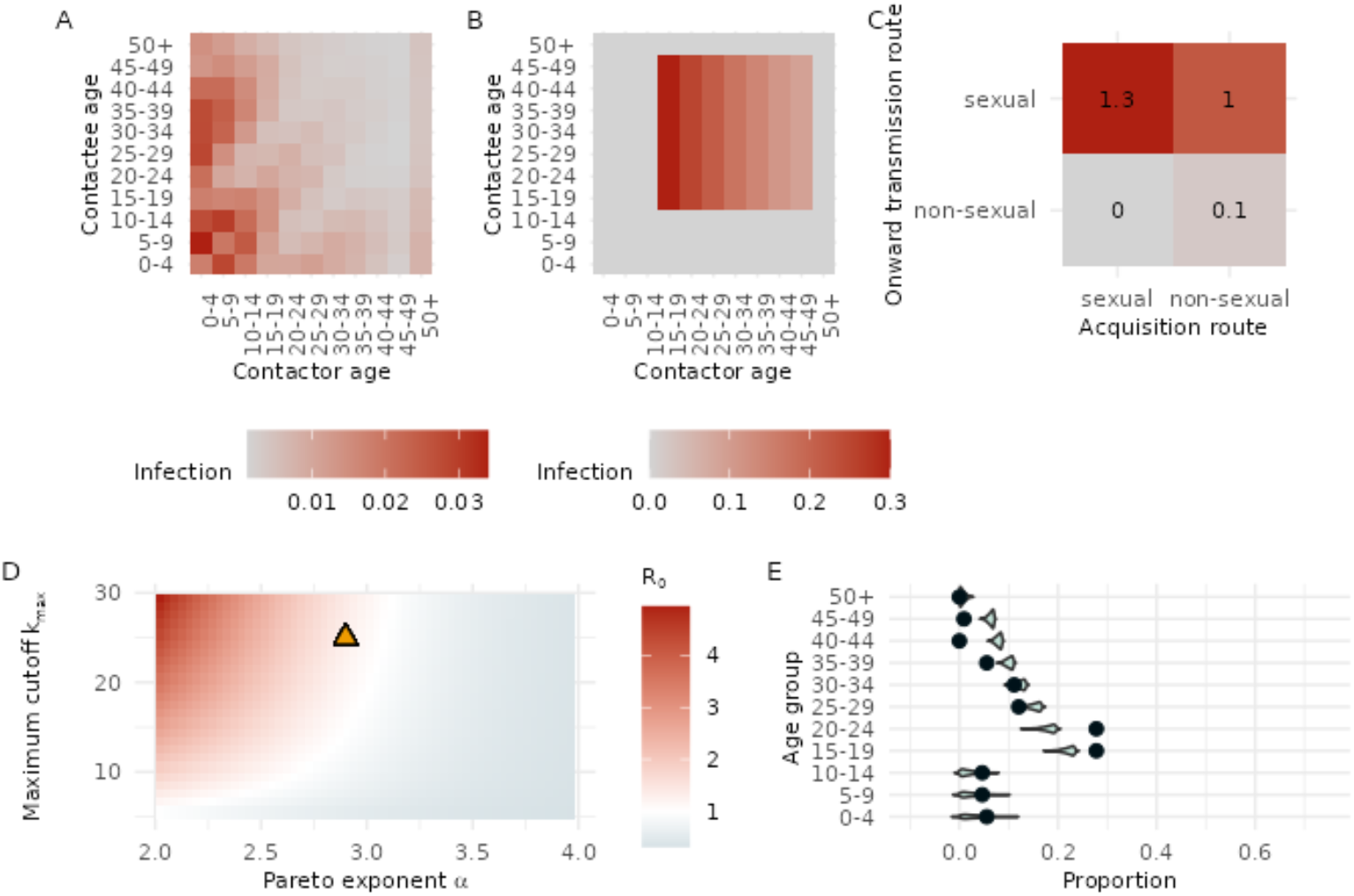
Projected next-generation matrices, reproduction numbers, and age distribution of infection. (A) Submatrix of the NGM representing transmission via non-sexual contacts from cases that were infected via non-sexual contact. (B) Submatrix of the NGM representing transmission via sexual contacts from cases aged 15 years and older who were infected via sexual contact. (C) Summarized route-to-route reproduction numbers. (D) Projected range of basic reproduction numbers as a function of the Pareto exponent (lower values indicate heavier-tailed degree distributions) and the maximum cutoff (maximum number of sexual partners). Yellow triangle indicates the baseline scenario. (E) Projected age distribution of infection incidence (blue density plots) compared to observed case data (black dots) from Kamituga, South Kivu province, the Democratic Republic of the Congo, between October 2023 to March 2024.

In addition to the baseline scenario, we accounted for the presumed uncertainty in the current estimates of parameters concerning the infectious period and SARs for clade Ib MPXV, and parameters around sexual behaviour. We projected route-to-route-specific reproduction numbers across the possible parameter range using Latin hypercube sampling (33) (see **Methods** for details on the explored parameter spaces). The projected values largely varied by route of transmission, ranging from 0.12-1.45 for *R*_*s,s*_, 0.28-1.11 for *R*_*s,ns*_, 0-0.07 for *R*_*ns,s*_, and 0-0.25 for *R*_*ns,ns*_. The projected basic reproduction number, *R*_0_, was 1.31 for the baseline scenario and ranged from 0.15 to 1.48. Our analysis revealed that the basic reproduction number *R*_0_ was primarily determined by *R*_*s,s*_ and *R*_*s,ns*_, the potentials for sexual-to-sexual and sexual-to-non-sexual transmission, respectively, and was sensitive to the change in the assumed distribution of the number of sexual partners (degree) (**Figure 1(D)**). See **Figure S1** in **Supplementary Materials** for the change in *R*_*s,s*_ and *R*_*s,ns*_ with respect to the assumed distribution. A higher maximum number of partners an individual can have at a time, and a heavier-tailed degree distribution led to an increased overall transmission potential *R*_0_. Our derivation of the NGM elements also suggested that the expected number of secondary infections caused by transmission via sexual contact is determined by the first and second moments of the degree distribution (see **Supplementary Materials**). This means that we can project the transmission potentials accurately if the mean and variance of the degree distribution are available, without knowing the entire degree distribution.

We projected a normalized age distribution of incidence of infection during an exponential growth phase by using the NGM, and compared it to the observed age distribution of clade Ib cases in Kamituga between October 2023 to March 2024 (1) (see **Methods** for details on the computational procedure). The projected age distribution closely aligned with the observed data, despite the wide range of parameter sets explored, indicating empirical support for the model structure used in this study (**Figure 1(E)**). The relative incidence among late teens and early twenties was higher in the observed data, likely owing to how the actual outbreak started with cases that belonged to those age groups in Kamituga.

### Time-varying contribution to transmission dynamics by route of transmission and age group

We projected the time-varying contribution of each route of transmission and age group, both by simulating an epidemic and by analytically deriving the time-varying NGM of the model (see **Supplementary Materials**). First, the projected epidemic trajectories under the baseline scenario are presented in **Figure 2 (A)**. This result shows that transmission via sexual contacts dominates the dynamics in the initial phase of an epidemic in terms of higher relative incidence compared to transmission via non-sexual contacts, but later transmission via non-sexual contacts becomes more prominent, leading to higher total incidence of infection. In this scenario, the time-varying reproduction number *R*(*t*) (computed as the dominant eigenvalue of the time-varying NGM, see **Supplementary Materials**) was 1.3 at the beginning (the projected *R*_0_) and started declining rapidly as transmission via sexual contacts becomes less dominant. In the later phase, *R*(*t*) declined to an asymptote of 0.45 due to the accumulated immunity among sexually active groups, leading to a self-contained outbreak. We performed a sensitivity analysis with the community contact matrix that includes all non-household contacts, which showed that the shift in transmission from sexual to non-sexual contacts occurred earlier due to the higher transmission rate via non-sexual contacts (**Figure S2** in **Supplementary Materials**).

**Figure 2.**
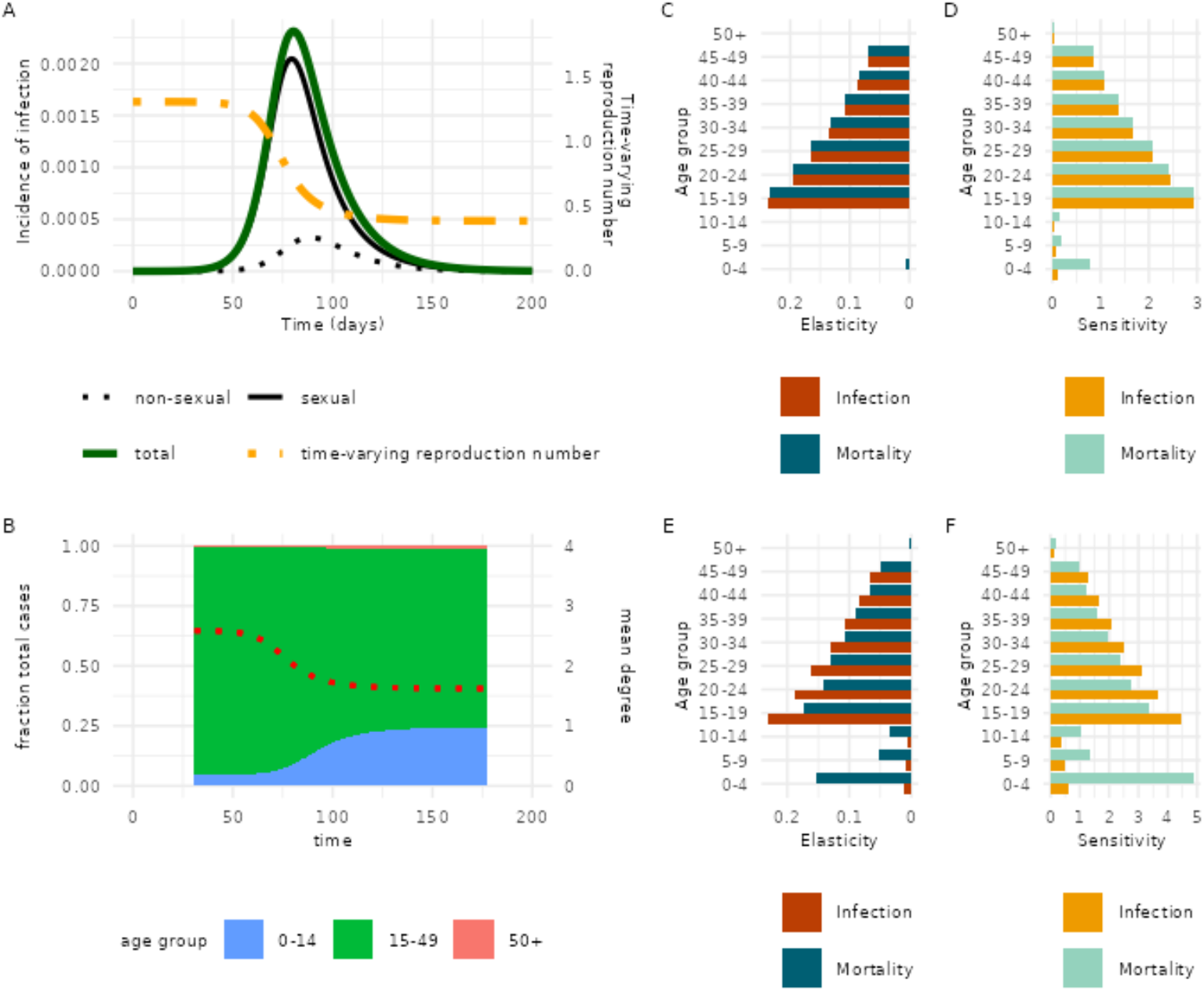
Projected epidemic dynamics over time by transmission route and age group. (A) Projected incidence of infections per day by transmission route shown alongside the corresponding time-varying reproduction number. (B) Age distribution of incidence of infection over time, shown alongside the mean number of sexual partners (red dotted line). (C, D) Elasticity and sensitivity of the next-generation matrix (NGM) and mortality-weighted NGM at the early stages of the epidemic. (E, F) Sensitivity and elasticity metrics at a later stage (day 200) of the epidemic.

In real-world outbreaks, the underlying dynamics cannot be directly observed. The developed network epidemic model offers two ways of assessing the epidemic phase over time using observable epidemiological data: the time-varying age distribution of relative incidence, and the time-varying mean degree of cases (**Figure 2(B)**). Our analysis shows that the age distribution of relative incidence is concentrated among sexually active age groups when transmission via sexual contacts dominates the incidence. As its contribution declines over the course of the epidemic, the relative incidence increases among the younger, sexually inactive age groups. The declining contribution of transmission via sexual contacts can also be measured by monitoring the degree of cases; in the baseline projection, the mean degree of newly infected sexually active individuals starts at 2.6 at the beginning and rapidly declines to an asymptote of 1.6. This reflects that individuals infected early in the epidemic typically have a higher number of partners than those infected later, when transmission via sexual contacts contribute less to the dynamics.

We use targeted vaccination as an illustrative example to identify subgroups with the highest expected contribution to two epidemic outcomes: the number of secondary infections after one generation of transmission (i.e., the reproduction number) and its consequent overall mortality. These contributions were measured in terms of cumulative sensitivity and elasticity, quantifying the relative and absolute contributions of each age group to infection and mortality. The analysis is based on perturbing the (time-varying) NGM and mortality-weighted NGM by immunizing one susceptible individual in a given group, as would occur through vaccination (see **Methods** and **Supplementary Materials**). We present the quantified age-group- and outcome-specific contributions at the initial stages of an epidemic (**Figure 2(C)(D)**) and at day 200 (**Figure 2 (E)(F)**). In the initial stages of the epidemic, younger adults aged 15-24 years contribute to both epidemic outcomes the most, making them the optimal target group for interventions. However, at day 200, the relative contribution to onward transmission declined among sexually active age groups, and the groups contributing to the overall mortality most were young adults aged 15-19 years and children aged 0-4 years (**Figure 2 (E)(F)**). Consequently, at day 200 after the initial outbreak, targeting both younger adults (15-19 years) and younger children (0-4 years) would be the most effective to minimize the expected overall mortality. We found that higher relative contributions of 0-4 years to the overall mortality would arise around day 160-200 by computing the time-varying age-specific sensitivity and elasticity over time (**Figure S3** in **Supplementary Materials**). We performed a sensitivity analysis using an alternative mortality-weighting approach based on crude CFRs for clade Ib in South Kivu (3). This result indicated that the relative contribution of the 0–4 year age group to overall mortality was diminished compared to the main analysis, due to the substantially smaller differences in age-specific CFRs used in the sensitivity analysis (**Figure S4** in the **Supplementary Materials**).

### Effect of transmission route on epidemic size

To examine how increased transmissibility via sexual and non-sexual contacts affects the epidemic size (fraction of the population that is ultimately infected if no interventions were taken), we projected epidemic trajectories and cumulative infections under different parameter scenarios by varying SARs for each transmission route (**Table 1**). Four alternative scenarios were explored by lowering SAR via sexual contacts and increasing SAR within households (household SAR), while keeping the same *R*_0_ = 1.31 as the baseline. With the baseline scenario, the epidemic size was the smallest (9.1%), and transmission via sexual contacts consisted of 84% of the total infections. In scenarios with higher household SAR, the epidemic size increased, driven by the increased proportion of transmission via non-sexual contacts interacting with transmission via sexual contacts (*R*_*ns,s*_). Notably, a moderate increase in the value for non-sexual-to-non-sexual transmission of *R*_*ns,ns*_ from 0.11 to 0.67 was sufficient to shift the dominant route of transmission from sexual to non-sexual contacts, resulting in a decrease from 89% to 41% of total infections caused by transmission via sexual contacts (comparing baseline to scenario B). This ‘switching’ dynamics aligns with a theoretical prediction from a previous study (23) and is particularly relevant because this moderate value for non-sexual-to-non-sexual transmission of *R*_*ns,ns*_ around 0.4−0.6 overlaps with the historical estimates of reproduction numbers for clade Ia MPXV (34, 35) and the assumed household SAR used to determine *R*_*ns,ns*_ was also comparable to the values reported in previous (non-clade Ib) mpox outbreaks (36). Moreover, the interaction between the two routes of transmission is crucial in the epidemic outbreak: in scenario C, where the reproduction number of each of the four submatrices of the NGM are less than one, an outbreak can still occur (*R*_0_ = 1.31), leading to a larger epidemic size than in the baseline scenario. In all alternative scenarios, larger epidemic sizes were projected with greater household SAR, even though the values of *R*_*ns,ns*_ were smaller than or closer to 1. Larger epidemic sizes were driven by the earlier shift in the dominant relative incidence from transmission via sexual contacts to transmission via non-sexual contacts (see the projected epidemic curves for each scenario in **Figure S5**).

**Table 1.**
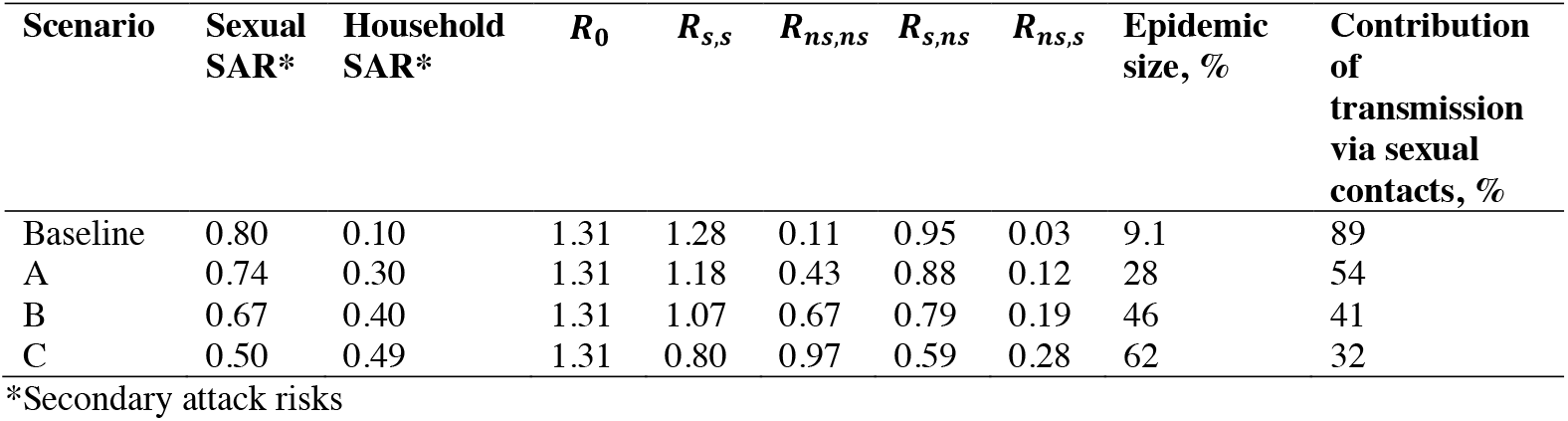
Epidemic size and contribution of sexual transmission under varying SARs. Epidemic size and percentage contribution of transmission via sexual contacts to the total epidemic size are shown across scenarios with different assumed SARs for transmission via sexual (*s*) and non-sexual (*ns*) contacts, while maintaining a constant basic reproduction number (*R*_0_ = 1.31). In scenario C, the household SAR is set to 0.49 (instead of 0.50) to ensure that all reproduction numbers remain below one.

| Scenario | Sexual SAR* | Household SAR* | $R_0$ | $R_{s,s}$ | $R_{ns,ns}$ | $R_{s,ns}$ | $R_{ns,s}$ | Epidemic size, % | Contribution of transmission via sexual contacts, % |
| --- | --- | --- | --- | --- | --- | --- | --- | --- | --- |
| Baseline | 0.80 | 0.10 | 1.31 | 1.28 | 0.11 | 0.95 | 0.03 | 9.1 | 89 |
| A | 0.74 | 0.30 | 1.31 | 1.18 | 0.43 | 0.88 | 0.12 | 28 | 54 |
| B | 0.67 | 0.40 | 1.31 | 1.07 | 0.67 | 0.79 | 0.19 | 46 | 41 |
| C | 0.50 | 0.49 | 1.31 | 0.80 | 0.97 | 0.59 | 0.28 | 62 | 32 |

\*Secondary attack risks

## Discussion

Our results suggest distinct epidemic dynamics of clade Ib MPXV may be driven by the coexistence of two transmission routes. Importantly, the dominant route of transmission may shift over time from transmission via sexual contacts to non-sexual contacts, depending on their relative transmission potentials. Such shifts may be detected using observable proxies, such as change in the age distribution of cases or in the mean number of sexual partners among confirmed cases, when measured consistently over time. If transmission via sexual contacts swiftly declines but transmission via non-sexual contacts continues over a prolonged period, that may indicate a higher transmission potential via close physical contacts in households or communities, leading to larger epidemic sizes. Monitoring such switching dynamics is crucial for anticipating the potential outcome of long-term epidemics and for informing response strategies in regions that may face future introductions of clade Ib MPXV.

Optimal target groups for intervention would differ depending on the epidemic outcome to minimize, as well as the timing of intervention. In our projections, secondary infections via non-sexual contacts were primarily associated with young children, whereas those via sexual contacts were concentrated among younger adults. Elasticity analyses indicated that targeting sexually active age groups, especially those aged 15-24 years, would be the most effective intervention to reduce both infections and mortality in the initial phase of an epidemic. However, once an epidemic progresses and immunity accumulates, the effectiveness of such intervention strategies on sexually active age groups would be attenuated. If increasing incidence rates among young children are observed, this may imply that sexually active groups may no longer be the optimal target groups for intervention. In contrast, those groups may remain relevant for outbreak responses in countries or regions that are still in their initial phase or have not yet experienced mpox outbreaks. These considerations are important for planning response strategies for ongoing and potential outbreaks, where anticipating the risk of sustained outbreaks following importation events is essential.

Our analyses underscore two critical data gaps: the SARs for each transmission route, and the sexual partnership distribution over relevant timeframes, particularly from the affected settings. Sexual behavior data are often reported on annual scales (37), which may not reflect the sexual contacts during the much shorter time scales for the typical infectious period of MPXV (38, 39). The simulated ranges of the reproduction number *R*_0_, the potential for sexual-to-sexual transmission (*R*_*ss*_), and sexual-to-non-sexual transmission (*R*_*s,ns*_) under varying degree distributions for the number of sexual partners highlighted that the misspecification of the degree distribution can lead to incorrect estimates of reproduction numbers. Our analysis also showed that modest increases in SARs for transmission via non-sexual contacts could be sufficient to shift the dominant transmission route and increase the epidemic size. These insights are essential to anticipate further international spread of clade Ib MPXV beyond the currently affected population groups and countries.

There are several limitations in this study. First, there are simplifying assumptions in our model; we assumed that both transmission routes had the same underlying generation time, although recent studies reported that the realized generation time was shorter for transmission via sexual contacts than that of transmission via non-sexual contacts (8). Our model structure is based on a closed population and does not incorporate spatial transmission or importations of cases. It also does not account for sex-specific differences, despite the fact that female sex workers were the initially affected population in Kamituga (1, 11) or differing age-sex distributions in sexual contacts (40). Second, contact matrices and the degree distribution used in this study may not reflect the actual contact patterns during the outbreak in Kamituga, for instance, due to a precautionary response as seen in the 2022 global outbreak (41). One-time sexual contacts, such as transactional sex, are not explicitly modeled and assumed to be represented by higher degrees of the partnership distribution. Empirical data are needed to determine the sexual partnership distribution, the age at the start of sexual activity (42), and the sex- and age-dependency of sexual behaviors when applying the proposed framework. Third, our analysis does not account for pre-existing or heterogeneous immunological status in the population, such as partial immunity among older adults due to historical smallpox vaccinations (43). The proposed approach could be further refined by incorporating such conditions, including nutritional and immunocompromised status (40), which may affect susceptibility and disease severity in target groups. Lastly, the quantities presented in this study should not be interpreted as empirical estimates, but rather as model-projected values, conditional on assumed scenarios. We focus on projecting potential epidemic outcomes under a large range of plausible conditions, rather than reconstructing the specific dynamics in Kamituga or inferring epidemic characteristics such as the duration of the outbreak, which is more prolonged in the actual outbreak. While model calibration to case data from Kamituga is theoretically feasible, such inferences would require careful interpretation, given the limitations imposed by armed conflict and limited surveillance infrastructure (44).

Our model can be applied to support risk assessments of ongoing mpox outbreaks, including in Kinshasa where clade Ia and Ib are co-circulating (45) and the new clade IIb outbreak in Sierra Leone (16); in both places, adults aged 16 to 35 years have been the most affected age groups. This study also contributes to the advance in modelling emerging infections with multiple, interacting transmission routes within age-structured populations with a network structure. Our mathematical analysis and projected epidemic sizes demonstrate that interactions between different transmission routes play a crucial role in shaping transmission dynamics, and that discarding them may lead to completely different epidemic outcomes. The proposed approach links qualitative understanding about transmission dynamics with empirical data, and provides a simple principle for identifying optimal subgroups for targeted interventions given available observations.

In conclusion, our findings highlight the critical need to adapt intervention strategies to the evolving dynamics of mpox outbreaks. In the presence of multiple transmission routes, models that explicitly capture their coexistence and interaction, such as the one proposed here, are essential for anticipating future epidemic trajectories. Optimal control strategies vary over time, depending not only on the epidemic phase but also on the specific outcome to be minimized, whether it be total infection or mortality. For countries that are at risk for importing new mpox cases, this highlights that continuous monitoring remains crucial to detect shifts in the dominant route of transmission and to support timely, effective public health responses.

## Materials and methods

### Two-level network epidemic model

The network epidemic model consists of two contact layers to represent transmission via sexual and non-sexual contacts. Sexual contacts occur among sexually active individuals within fixed partnerships that together form a static sexual network. This network is constructed using a configuration model to match a prescribed degree distribution (46), where the degree represents the number of sexual partners over the time window of a typical infectious period, under the assumption that individuals do not change their partners over the course of the epidemic. Non-sexual contacts occur across the entire population for all individuals through age-dependent random mixing, represented by a contact matrix whose elements denote the average number of daily contacts per individual between and within age groups.

The two contact layers are connected to each other through individuals who are present in both layers. Superimposed on the population with the two contact patterns is a compartmental SEIR infection. Transmission can occur via either sexual or non-sexual contacts, depending on the route of contact between an infectious and a susceptible individual. The transmission probability per sexual partnership is the same and independent of the total number of sexual partners of the individuals in the partnership. The model is deterministic in nature, and can be fully captured by three sets of parameters: i) degree distribution relating to sexual contacts, ii) contact matrix relating to non-sexual contacts, iii) infection parameters. Moreover, we focus on projections of transmission potential and epidemic trajectories over a period of less than one year; we ignore aging or demographic turnover (e.g., migration, births and natural deaths) as well as the waning and the pre-existence of immunity. The network epidemic model can be described by a set of ordinary differential equations (ODE), and the full description is provided in **Supplementary Materials**.

### Next-generation matrices and reproduction numbers

To quantify the age-specific contribution to transmission, we first introduce the next-generation matrix ***K***, which contains as its elements the number of secondary infections by age group and transmission route. By summarizing all elements, the full NGM ***K*** can be expressed as:

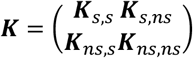

where the four block submatrices can be viewed as the NGMs that describe the secondary transmission from one route to another or within the same route in the absence of other route-to-route transmissions: ***K***_***s***,***s***_ (transmission from sexual to sexual contacts), ***K***_***s***,***ns***_ (from non-sexual to sexual), ***K***_***ns***,***s***_ (from sexual to non-sexual), ***K***_***ns***,***ns***_ (from non-sexual to non-sexual). Each submatrix is a M x M matrix with M the number of age groups and consists of the elements of the age-specific number of secondary infections caused via one route of transmission by a case infected via a specific route of transmission. We derive all NGMs from the network epidemic model developed above, and detailed derivations and their interpretations are provided in the **Supplementary Materials**.

We then characterize the route-to-route-specific transmission potential and the overall dynamics by computing distinctive reproduction numbers. The dominant eigenvalue of the full NGM at time 0 (i.e., the time at which all individuals are susceptible) is the basic reproduction number *R*_0_, determining whether a major outbreak may occur or not (29, 32). Route-specific reproduction numbers can be computed by taking the dominant eigenvalue of the four submatrices defined above, and therefore we can obtain four quantities: *R*_*s,s*_ (from sexual to sexual), *R*_*s,ns*_ (from non-sexual to sexual), *R*_*ns,s*_ (from sexual to non-sexual), *R*_*ns,ns*_ (from non-sexual to non-sexual). Note that these reproduction numbers are derived with an implicit assumption that there are no interactions between transmission routes due to the block-wise decoupling of the full NGM. Therefore, they should be interpreted as the average number of secondary infections within each route pair with an assumption that no cross-transmission occurs between routes. To determine *R*_0_ the full NGM ***K*** is needed. By further extending this next-generation approach, one can derive time-varying NGMs and compute the time-varying reproduction number *R*(*t*) (**Supplementary Materials**), and use *R*(*t*) to analyze the evolving overall transmission potential.

### Perturbation analysis of next-generation matrices

By analyzing the constructed NGMs at different points in time, we can determine the age group that contributes most to onward transmission at each time point. Such age-specific contributions can be measured by two indicators: cumulative sensitivity 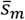 and cumulative elasticity *ē*_*m*_ (for age group *m*). Sensitivity measures the absolute change (hereafter *R*, dropping *t* for simplifying notation) with respect to a small change in an element of a NGM (i.e., 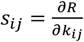). Elasticity measures the proportional (relative) change in response to a proportional (relative) change in an element of a NGM, and is expressed by 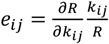. Cumulative sensitivity and elasticity quantify the total impact of a small (absolute and relative) change in a single targeted age group *i* on *R*, and they are given by 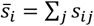 and *ē*_*i*_ = ∑_*j*_ *e*_*ij*_, respectively (29, 47).

The perturbation analysis described above can be applied to other projection matrices (30, 48). We constructed a projection matrix for death due to secondary infections at time *t* by multiplying a matrix of age-specific infection mortality ratio:

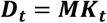

where ***M*** is a diagonal matrix with the elements of age-specific infection mortality ratios, and ***K***_***t***_ is the (full) NGM at time *t*. By quantifying age-specific cumulative sensitivities or elasticities, we can determine the group that contributes most to the mortality due to secondary infections caused by that group. This approach is extendable to other disease outcomes or indicators (e.g., hospitalizations, or DALYs) as long as the outcome can be expressed as a multiplicative effect, as demonstrated in (30). For the infection mortality matrix ***M***, we used the pooled CFR estimates of clade I reported by (10) as a proxy in the main analysis. As a sensitivity analysis, we also used the crude CFR values for South Kivu reported by WHO (3).

### Parameterization of contact networks

In our network epidemic model, age-specific non-sexual contacts were parameterized by using synthetic contact matrices estimated for the DRC by Prem et al. (49). Contacts in the contact matrices are defined as conversational and/or physical contacts as used by social contact surveys (50, 51). Contacts in the household matrix are assumed to largely represent individuals’ interactions with close physical proximity. We therefore used the household contact matrix for the main analysis and the contact matrix that covering contacts in all settings (i.e., contacts within and outside the household, including contacts in a community) for sensitivity analysis (**Figure S6**). While the contact matrices do not distinguish contact types by conversational or physical contact, it is likely that non-household settings include a higher proportion of non-physical (i.e., conversational-only) contacts. Therefore, the projections in the sensitivity analysis should be interpreted as upper-bound estimates under this assumption as our use of the all-settings matrix may overestimate the contact rates for transmission via non-sexual contacts.

To represent the sexual contact network, we used a degree distribution that represents the number of sexual partners in the time window of a typical infectious period. We used a truncated power-law degree distribution given by:

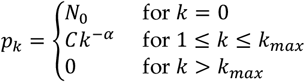

where *N*_0_ is the proportion of individuals with zero partners, *α* is the Pareto exponent, *C* is a normalization constant, and *k*_*max*_ is the maximum degree. The same formulation is applied in e.g. (23). The truncated power-law distribution allows for empirical parameterization of the proportion of degree zero, as well as the maximum cutoff. As a baseline parameter set to visualize the main results, we used *N*_0_ = 0.14 based on a DHS survey in the DRC (40), *α* = 2.9 by referring to an estimated value for men in a sexual behaviour survey in Burkina Faso (Burkina Faso permits legal polygamy and has documented the presence of transactional sex work, similar to the DRC) (52), *k*_*max*_ = 25 to reflect the mean number of clients per female sexworker over infectious period of 10 days by linearly rescaling the observed number of clients over a week (53). Although the baseline parameter values and the consequent mean number were similar to the observed data of the number of sexual partners (12), there are large uncertainties in each parameter. Moreover, a broad range of parameter values could also be used for assessing how the outbreaks may develop in other countries that have different epidemiological and behavioral parameters than those of the DRC. Therefore, we took a range of each parameter, rather than using point estimates, when comparing the model-projection to the observed age distribution of incidence of infection using a Latin hypercube sampling (33) (see the summary of explored parameter ranges in **Table S1**). We assumed the same degree distribution for all sexually-active age groups (those aged 15−49 years, based on (40)). Age groups were defined as 0−4, 5−9, 10−14, …,40−44, 45−49, and 50+ years old, according to the age grouping in WHO reports (3).

### Epidemiological parameters

For the baseline scenario, we used the mean generation time of 10 days (based on the mean serial interval estimates of clade IIb in 2022 (54) and of clade Ib among cases infected with non-sexual contacts in eastern DRC (8)), latent period of 2 days (55), and infectious period of 8 days (56). Per-contact transmission rates for transmission via sexual and non-sexual contacts were specified by referring to the SAR observed in historical clade Ia outbreaks and clade IIb outbreaks, and household SAR of 0.10 and sexual SAR of 0.80 were used for the baseline scenario. We assumed the same per-contact transmission rates for both within-household and outside-household contacts as the focus of the sensitivity analysis of **Figure S2** was to check the impact of differential contact rates. Since both the natural history of clade Ib MPXV is still largely unknown, and for assessing how outbreaks may develop in other settings, we explored the wide range of the parameter space to project reproduction numbers and age distribution of incidence of infection. The explored range of the transmission parameters were summarized in **Table S1**, and its rationale is described in **Supplementary materials**.

When computing epidemic sizes under different scenarios (**Table 1**), the SAR of household transmission is varied, while keeping the basic reproduction number fixed to the baseline setting *R*_0_ = 1.31. Varying the SAR for household transmission fully determines the corresponding transmission parameter. The parameter of transmission via sexual contacts is determined through the basic reproduction number. As a consequence, also the SAR of transmission via sexual contacts is modified. Scenario C with a SAR for household transmission of 0.49 instead of 0.50 is chosen to ensure that *R*_*ns,ns*_ < 1.

### Projected age distribution of incidence of infection

The projected age distribution of incidence of infection was computed by taking the dominant (right) eigenvector of the NGM, based on its ergodic property (29, 48) The dominant right eigenvector converges to a stable distribution, which is proportional to the incidence of infection during the exponential growth phase. We summed the secondary infections from both transmission routes (sexual and non-sexual), computed the resulting age-specific incidence, and then normalized it to obtain the projected age-distribution of incidence. To assess convergence, we evaluated the damping ratio, defined as the ratio of the largest to the second-largest eigenvalue (48), indicating that the asymptotic distribution is reliably reached (with an error smaller than 1% of the dominant eigenvalue) after three generations of transmission.

### Outbreak data and demographic information

We collected the number of confirmed mpox cases in Kamituga, South Kivu province, the Democratic Republic of the Congo, between October 2023 and March 2024 (1). We aggregated sex to construct a (normalized) age distribution of incidence of infection. The nation-wide population age distribution in the DRC was retrieved from (57), assuming that the distribution is comparable in Kamituga to compare the observed incidence against the model-projected incidence.

## Data Availability

All codes and analyzed data are available on GitHub: https://github.com/rivm-syso/mpox-network

## Author contributions

Conceptualization: FM, KYL.

Data curation: FM, KYL.

Formal analysis: FM, KYL.

Investigation: FM, KYL.

Methodology: FM, KYL.

Software: FM, KYL, MvA.

Validation: FM, KYL.

Visualization: FM, KYL.

Writing – original draft: FM, KYL.

Writing – review & editing: FM, KYL, MX, MvA, NL, NH, EHV, JW.

## Data and code availability

All codes and analyzed data are available on GitHub: https://github.com/rivm-syso/mpox-network

## Supplementary Materials

**Figure S1.**
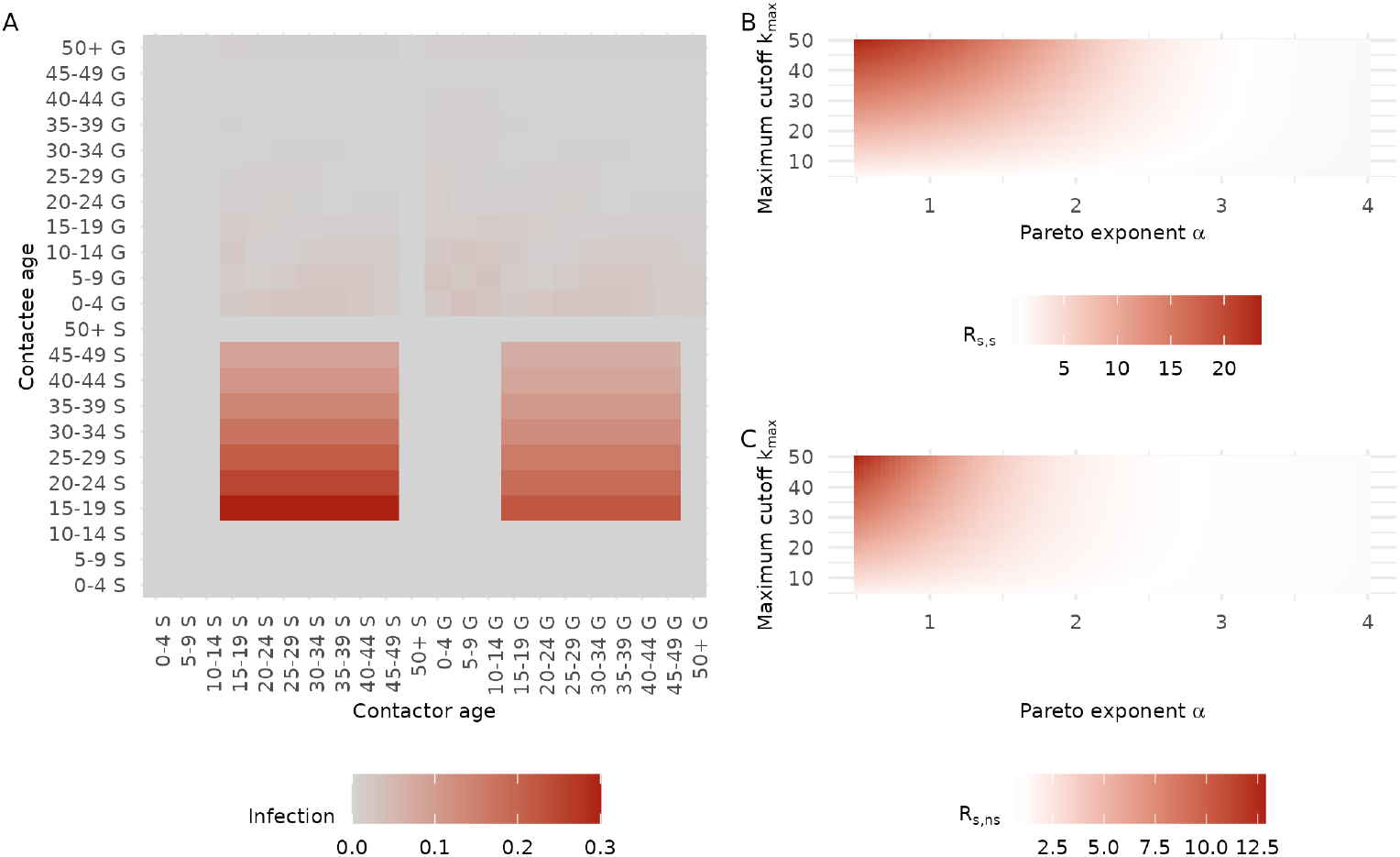
Projected next-generation matrix and reproduction numbers by transmission route and degree distribution parameters. (A) Full next-generation matrix (NGM) showing transmission via sexual and non-sexual (denoted with s and ns) contacts, stratified by age group. (B) Projected range of from-sexual-to-sexual reproduction numbers (*R*_*s,s*_) and (C) from-non-sexual-to-sexual reproduction numbers (*R*_*s,ns*_) as functions of the Pareto exponent (lower values indicate heavier-tailed degree distributions) and the maximum cutoff (i.e., the maximum number of sexual partners).

**Figure S2.**
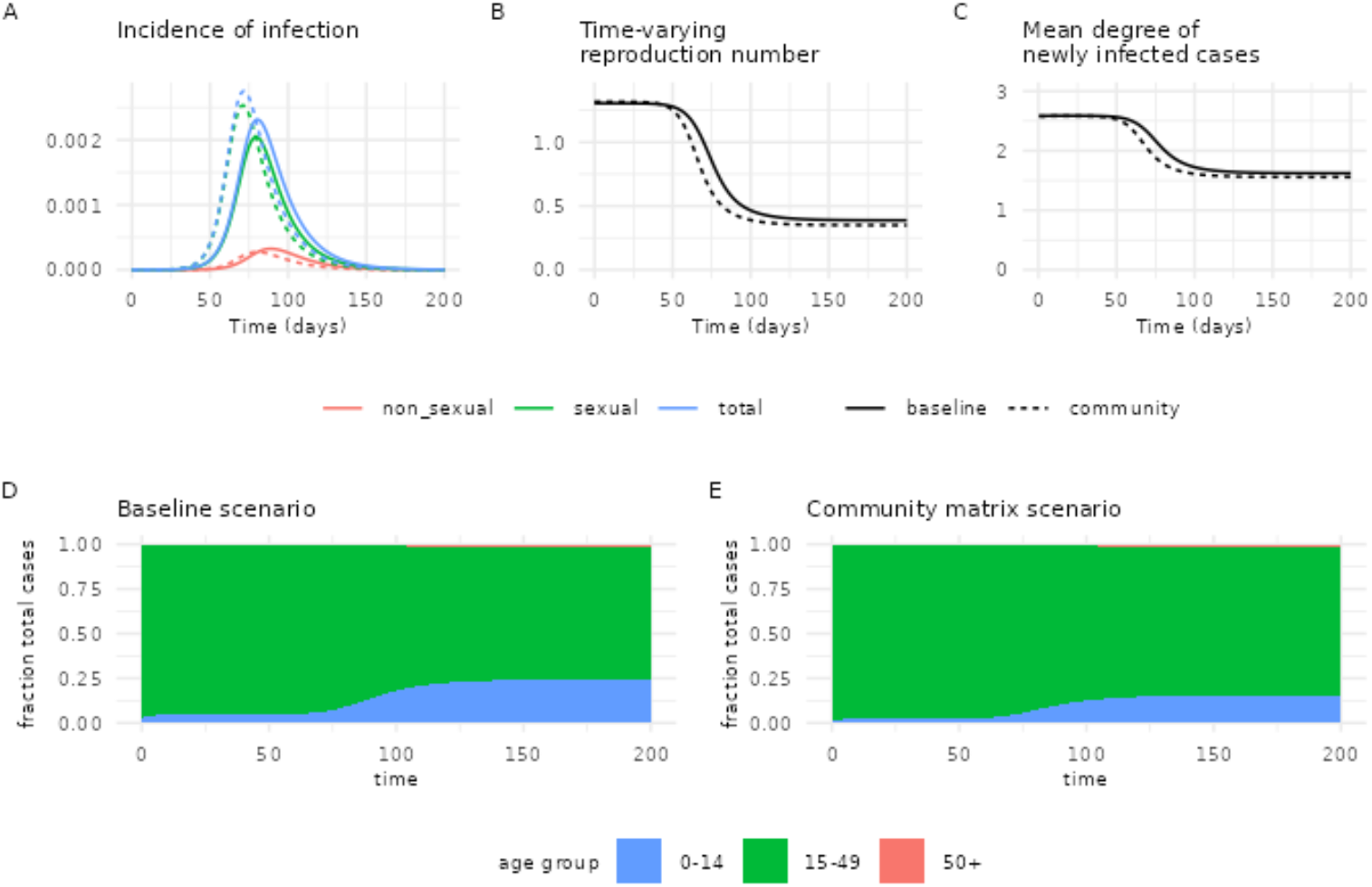
Comparison of baseline and community contact matrix scenarios. The baseline scenario uses a household contact matrix, and the community contact matrix scenario accounts for all contacts both within and outside households. (A) Incidence of infection per day by route of transmission. (B) Time-varying reproduction number. (C) Mean number of partners of a newly infected case (D, E) Age-distribution of infection over time for the baseline scenario (D) and the community matrix scenario (E).

**Figure S3.**
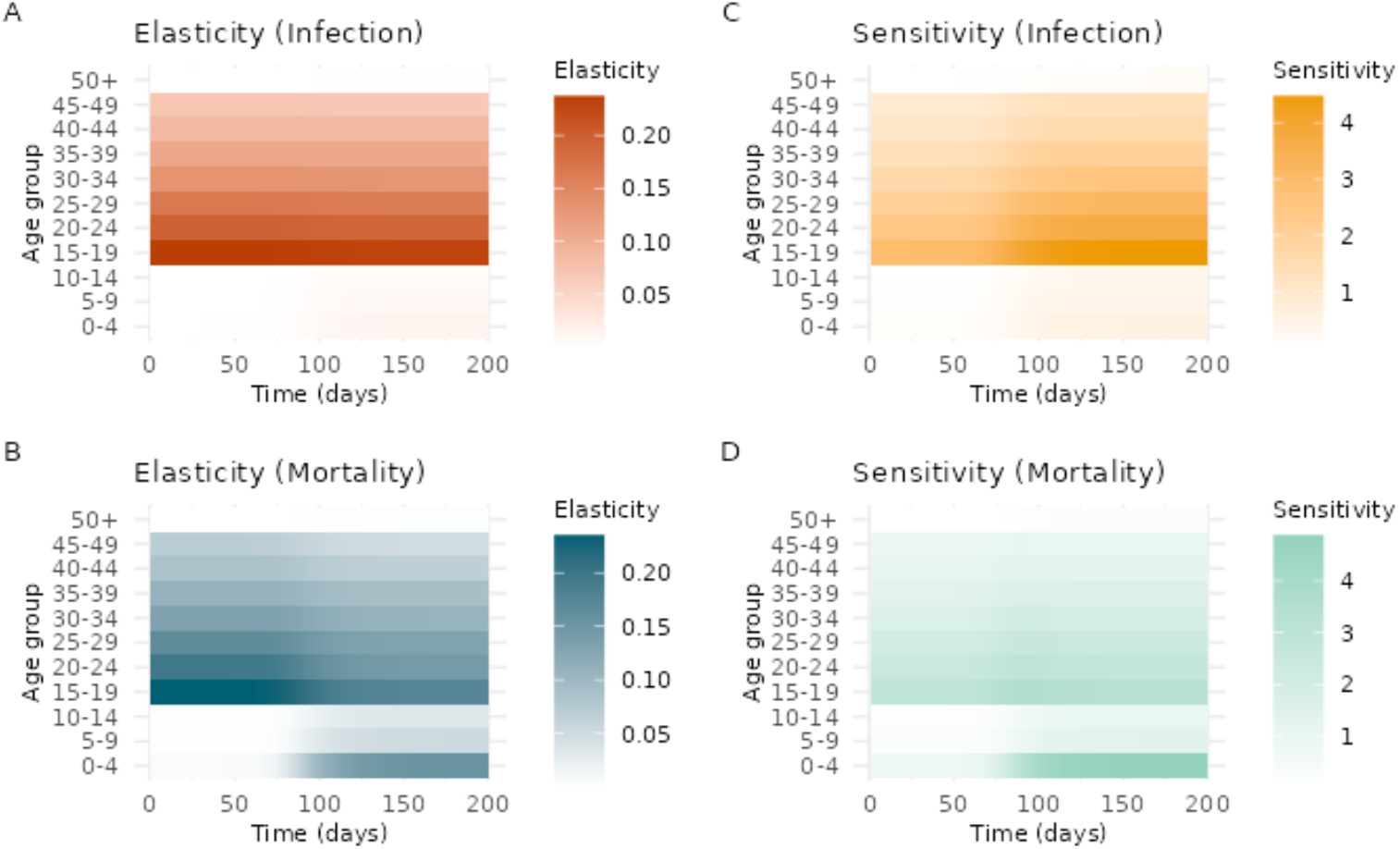
Projected age-specific sensitivity and elasticity for infection and mortality over time. Elasticity and sensitivity are calculated from the next-generation matrix (NGM) for infection and a mortality-weighted NGM for death. (A) Age-specific elasticity for infection. (B) Age-specific elasticity for mortality. (C) Age-specific sensitivity for infection. (D) Age-specific sensitivity for mortality. Elasticity and sensitivity values for each age group are to be compared relative to each other – higher elasticity or sensitivity means higher contribution of that age group. Compare to Figure 2 C-F in the main text, where elasticity (Fig. 2 C,E vs Fig. S3 A,B) and sensitivity (Fig. 2 D,F vs Fig. S3 C,D) for infection and mortality at time t=0 and t=200 are displayed.

**Figure S4.**
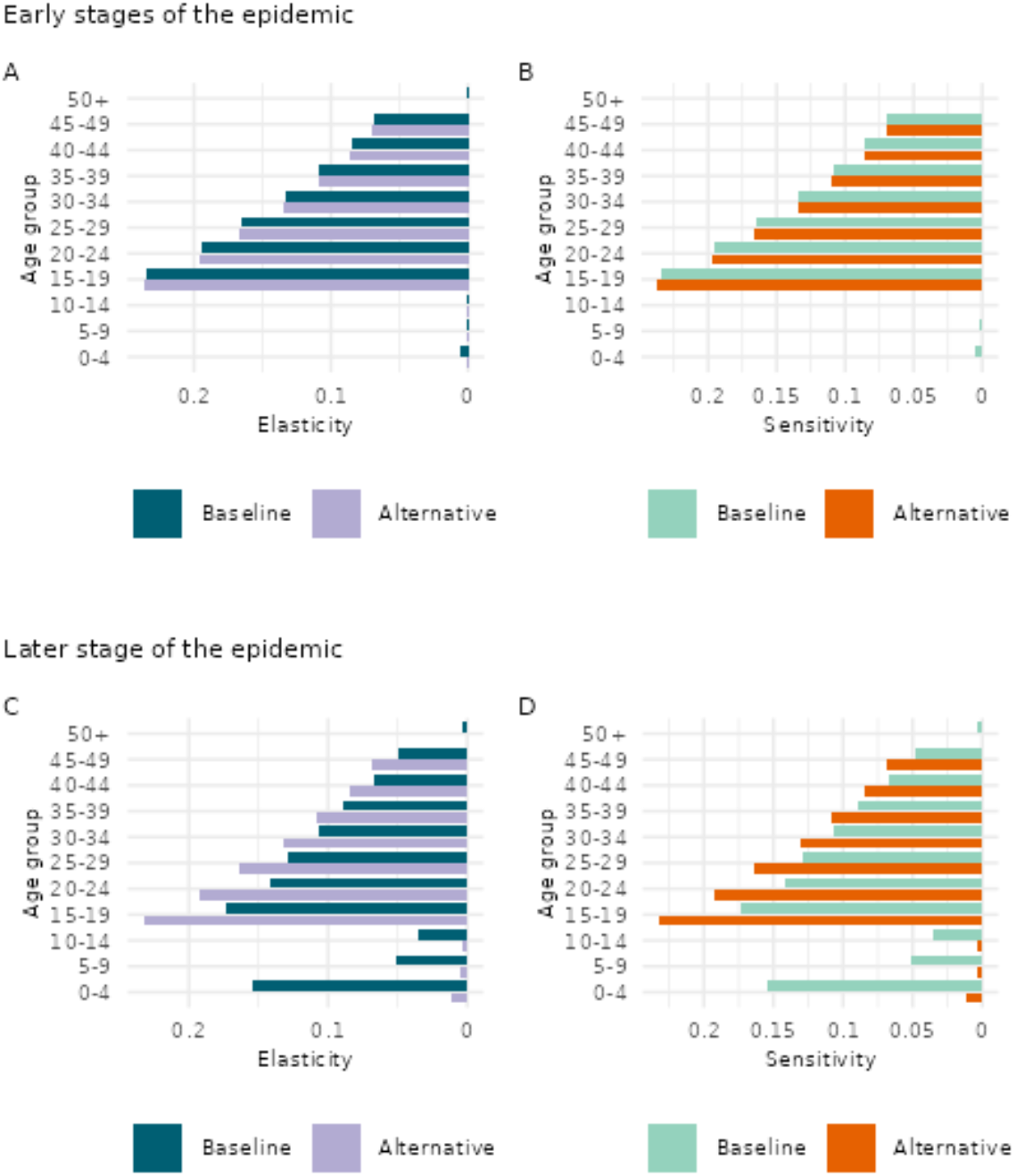
Comparison of the projected age-specific elasticity and sensitivity for mortality for the baseline mortality matrix and alternative mortality matrix. Elasticity and sensitivity are calculated from the mortality-weighted NGM for death. Alternative mortality matrix is based on the case fatality ratio in South Kivu reported by the Democratic Republic of Congo Ministry of Public Health (0.2% for 0-4 yo, 0.1% for 5-14 yo, and 0.2% for 15+) [1]. (A, B) Age-specific elasticity and sensitivity for mortality at the early stages of the epidemic (t = 0). (C, D) Age-specific elasticity and sensitivity for mortality at later stage of the epidemic (t = 200). Elasticity and sensitivity for the baseline mortality are also displayed in Figure 2 C-F in the main text.

**Figure S5.**
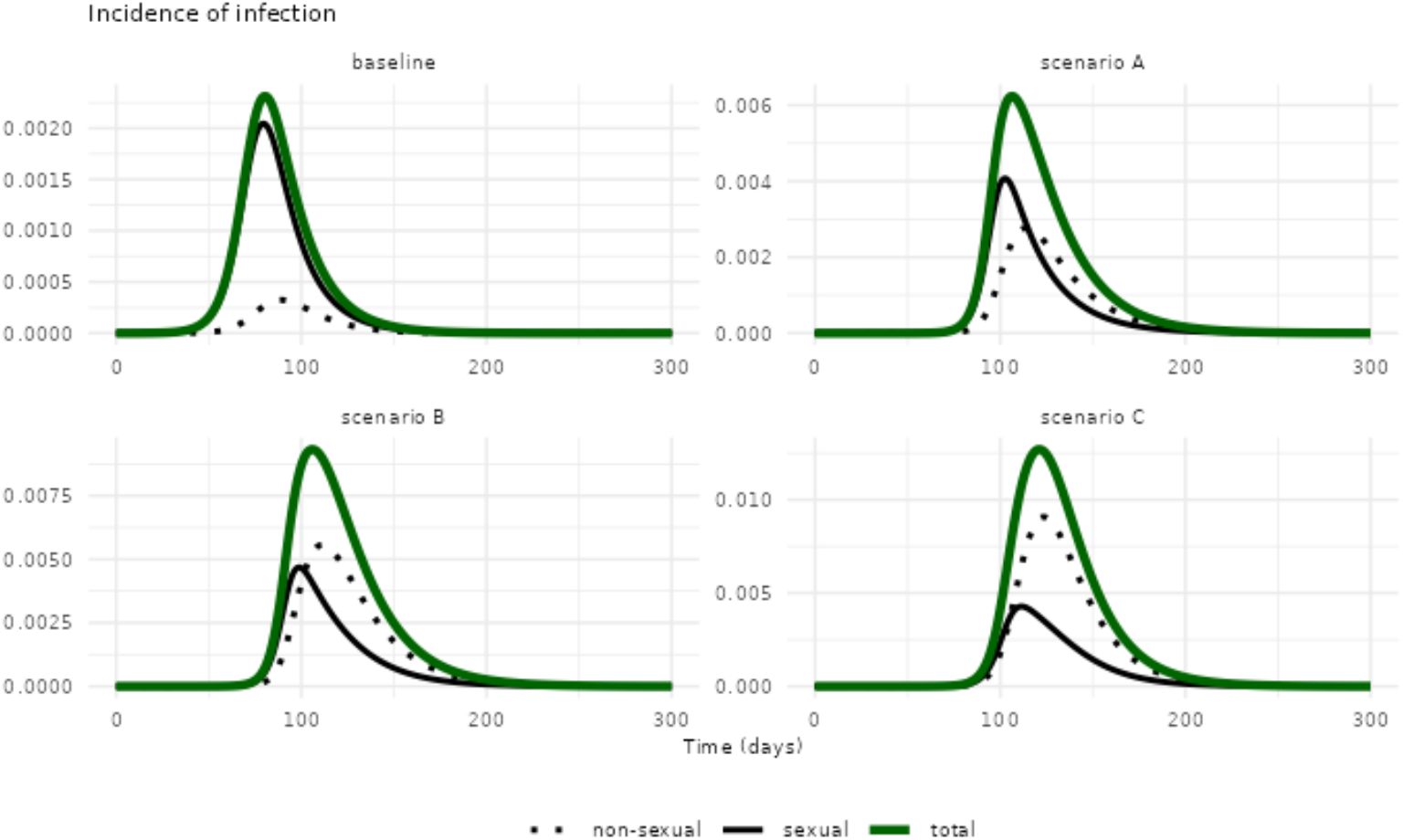
Projected incidence of infection by route of transmission. Comparing the baseline scenario with scenarios A, B and C where secondary attack risks of household and sexual contact are varied (refer to Table 1 in the main text).

**Figure S6.**
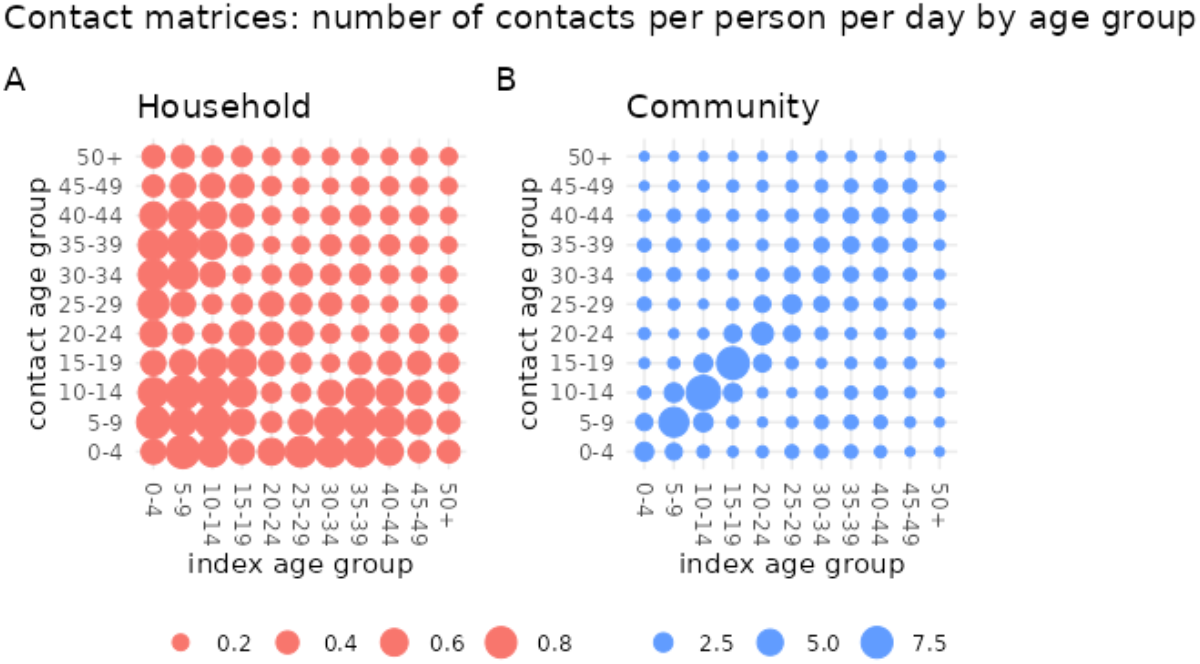
Synthetic contact matrices for the Democratic Republic of Congo. The number of contacts per person per day by age group in household setting (A) and in community (B). Household contact matrix is used for the baseline scenario, and the community contact matrix is used in the sensitivity analysis in Figure S2.

**Table S1.**
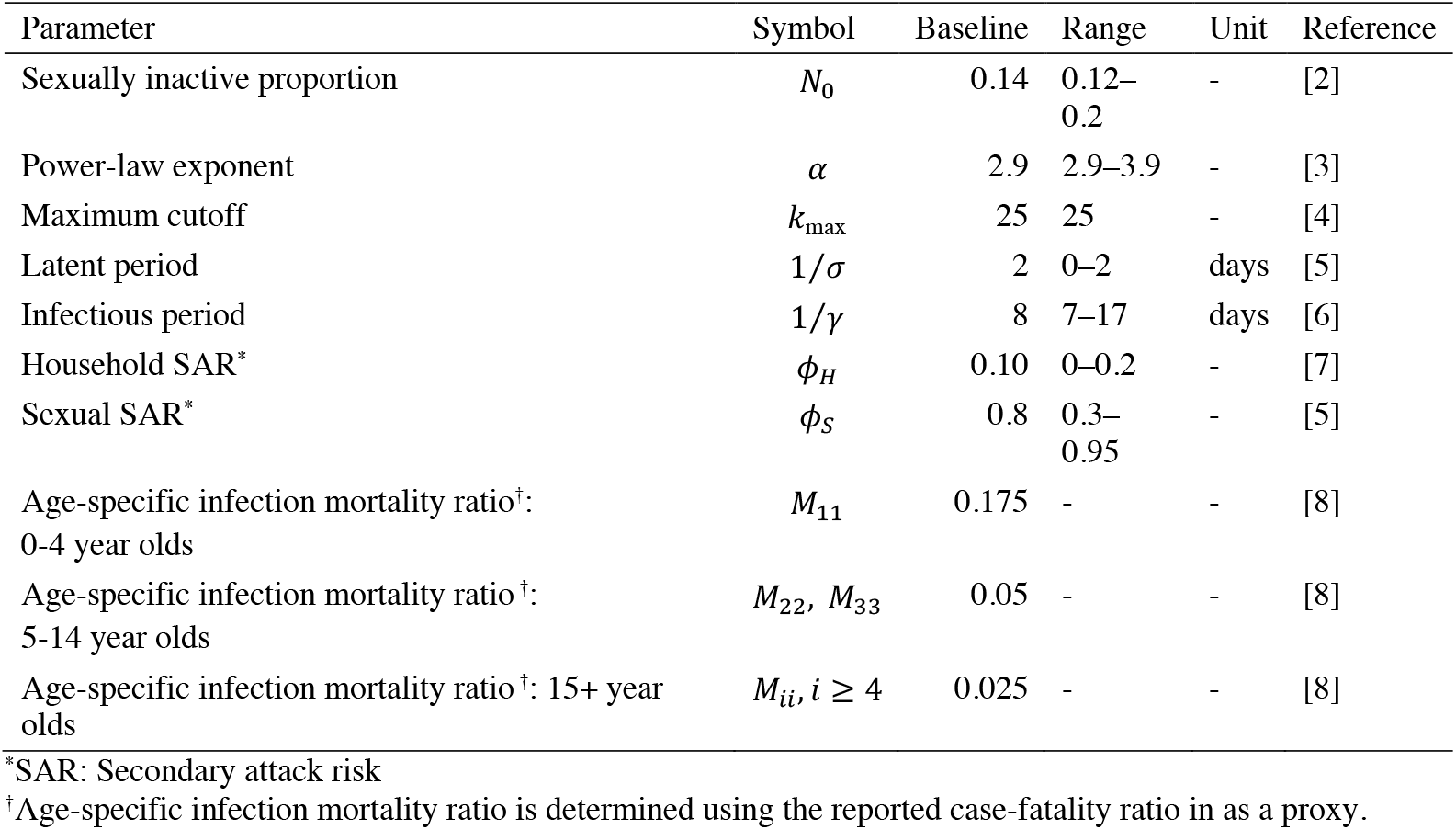
Baseline values and explored ranges of model parameters.

| Parameter | Symbol | Baseline | Range | Unit | Reference |
| --- | --- | --- | --- | --- | --- |
| Sexually inactive proportion | $N_0$ | 0.14 | 0.12–0.2 | - | [2] |
| Power-law exponent | $\alpha$ | 2.9 | 2.9–3.9 | - | [3] |
| Maximum cutoff | $k_{\max}$ | 25 | 25 | - | [4] |
| Latent period | $1/\sigma$ | 2 | 0–2 | days | [5] |
| Infectious period | $1/\gamma$ | 8 | 7–17 | days | [6] |
| Household SAR* | $\phi_H$ | 0.10 | 0–0.2 | - | [7] |
| Sexual SAR* | $\phi_S$ | 0.8 | 0.3–0.95 | - | [5] |
| Age-specific infection mortality ratio<sup>†</sup>:<br>0–4 year olds | $M_{11}$ | 0.175 | - | - | [8] |
| Age-specific infection mortality ratio<sup>†</sup>:<br>5–14 year olds | $M_{22}, M_{33}$ | 0.05 | - | - | [8] |
| Age-specific infection mortality ratio<sup>†</sup>: 15+ year<br>olds | $M_{ii}, i \geq 4$ | 0.025 | - | - | [8] |

\*SAR: Secondary attack risk
<sup>†</sup>Age-specific infection mortality ratio is determined using the reported case-fatality ratio in as a proxy.

### Supplementary text 1: Additional detail on parameter settings, model assumptions and scenarios considered

#### Explored ranges of epidemiological parameters

The parameter ranges were specified by referring to historically observed values: mean generation time of 7–17 days, mean latent period of 0–2 days, and mean infectious period of 7–17 days. The range of mean generation time reflected the shortest scenario referring to clade IIb MPXV outbreaks in 2022 [9] and the longest scenario based on historical estimates for clade Ia [10]. The shortest mean latent period was set as 0 days to represent possible infectiousness via sexual contact before experiencing recognizable symptoms. We referred to the reported range of household secondary attack risks (from 0 to 12%: 0-11% among countries in Africa [7], 12% in a historical outbreak in the DRC [11], and 7% in the US during the 2022 outbreak [12]).

#### Translating secondary attack risks into transmission rates

As the secondary attack risk per sexual partner can be computed as 1 minus the probability of escaping from all infectious contacts from a single sexual partner over the infectious period, the relationship between transmission rate β and the SAR of transmission via sexual contact *ϕ*_*S*_ is given by 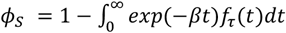 where *τ* is the infectious period with density function *f*_*τ*_. In our SEIR framework, 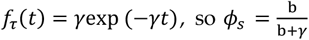. By rearranging this, the transmission rate for sexual contacts is given by 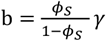. An implicit assumption here is that the number of contacts per partner per unit time is 1 (or, the contact frequency per day is involved in the transmission rate b). For the transmission rate in households or communities, we denote b_*H*_ by a household transmission rate, and *ϕ*_*H*_ the SAR of transmission within households. The household transmission rate satisfies 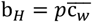 where 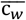 represents the average effective contacts per day within a household and *p* is the per-contact probability of infection. 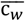 is computed by taking the dominant eigenvalue of age-weighted contact matrix 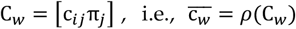. Using the same relationship between SAR and transmission rate as above, we obtain 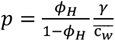. For the baseline scenario, the resulting per-contact probability of infection for non-sexual contacts is 0.025.

#### Additional model assumptions

In addition to the structural assumptions of the network model, we outline below several simplifying assumptions made when illustrating our modeling framework:

- No differences in sex in the model: we assume that contact rates (both sexual and non-sexual contact rates), the age distribution, and any other model parameters are not sex specific.
- Equal per-contact susceptibility and infectivity: we assume homogeneous susceptibility and infectivity across individuals, regardless of age and sex, due to limited data. That is, all individuals are equally likely to acquire and transmit infection given contact.
- No prior immunity existing in the population: the model assumes a fully susceptible population at the onset of the outbreak, reflecting the absence of prior immunity or protective effect from historical smallpox vaccination, which ended around 1980 in the DRC [13]. We opted for this conservative assumption given the limited information on cross-protection, waning immunity, and vaccination coverage in the affected populations.
- Vaccine efficacy and vaccine mode of action: when simulating targeted interventions with the sensitivity of NGMs, we consider the decrease of susceptible proportions among the targeted age groups. We assume that the vaccine confers protection against infection through an all-or-nothing mechanism (i.e., a proportion of vaccinated individuals are assumed to be fully protected from infection, while the remainder receives no protection). Vaccine efficacy (VE) is defined as the proportion of vaccinated individuals who are completely protected against infection, and VE against infection assumed to be 0.8 [14], [15].
- Initial conditions of numerical simulations: the model assumed that the epidemic is seeded by infectives in the Exposed stage (infected but not yet infectious) in the sexually active age group 20-24 years old who form a fraction 1/population size (i.e., 9 × 10^−6^) of the total population, which has a small effect at the very beginning of the epidemic only.

### Supplementary text 2: Mathematical details

#### 1 Model description

We consider a population that is structured according to age and sexual activity. Whether an individual belongs to the sexually active population or not depends on age (younger age groups are assumed to be sexually inactive). An individual who is part of the sexually active population has a certain (fixed) number of sexual partners *n* that is called the degree of the individual. The degree of an individual is determined by an age-*independent* degree distribution 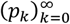, where *p*_*k*_ is the fraction of the population with degree *k, k*=0,1,2,… The network does not change in the period under consideration. A proportion of the sexually active population can be without partners, i.e. individuals with degree 0, where *p*_0_ > 0.

There are two types of contacts that can be made in the population. Individuals in the sexually active population can make (sexual) contact within their partnerships. On top of that, there are non-sexual contacts between any two individuals in the population according to the mass action principle. These non-sexual contacts are age-*dependent*. The sexual network is constructed by the configuration network construction [16]. We follow notation and terminology of [17]. In the configuration network construction, mathematically, an individual with degree *n* is in a sense a collection of *n* binding sites. Then binding sites are randomly paired up, and the corresponding individuals form a partnership with each other.

We consider a SEIR infection in the population with the sexual network and non-sexual contacts. Individuals are either *S* usceptible, *E* xposed to infection but not yet infectious, *I* nfectious, or *R*ecovered and immune from the infection for the duration of the epidemic. The latent period is exponentially distributed with mean 1/*σ*, and the infectious period is exponentially distributed with mean 1/*γ*. Latent and infectious period are independent of the route of transmission or the ages of the individuals involved. Transmission via sexual contact occurs at rate β from an infectious individual to each of his/her susceptible sexual partners, independently of the total number of partners. Transmission via non-sexual contact occurs at rate *pc*_*ij*_ from an infectious individual of age *i* to a susceptible individual of age *j*, where *c*_*ij*_ = *c*_*ji*_ is the per-pair non-sexual contact rate between individuals of age *i* and *j*, and *p* is the probability of transmission upon non-sexual contact between an infectious and susceptible individual.

The configuration network construction allows us to determine the state of an individual by considering the age and infection status of the individual. If the individual is sexually active, the state of an individual involves also the number of partners of the individual, and the age and infection status of each of these partners. By viewing sexually active individuals as collections of binding sites, we can reduce the description of the model to tracking susceptible binding sites, together with susceptible, exposed, and infectious individuals in the sexually inactive ages.

We let 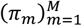 denote the population age distribution, where *π*_*m*_ is the proportion of individuals in age group *m*. The age groups *m* = 1, …, *N*_1_ − 1, *N*_2_ + 1, …, *M* comprise the sexually inactive population, and the age groups *N*_1_, …, *N*_2_ form the sexually active population, 1 ≤ *N*_1_ < *N*_2_ ≤ *M*. Consider a sexually active susceptible individual (i.e. age group *N*_1_ ≤ *i* ≤ *N*_2_) with degree *n* > 0, and examine one of his/her binding sites. Assume that the individual does not become infected through one of his/her *n* − 1 other binding sites for the period under consideration. Let *x*_(*i,n*),(*j,y*)_ denote the probability that a binding site belongs to a susceptible individual with age *i* and degree *n* and is occupied by an individual with age *j* and infection state *y, y* = *S, E, I, R*. Let *x*_*i*,0_ denote the probability an individual with age *i* and degree 0 is susceptible. Let *χ*_(*i,n*),(*j,k,S*)_ denote the probability that a binding site belongs to a susceptible individual with age *i* and degree *n* and is occupied by a susceptible individual with age *j* and degree *k*. Note that consistency requires 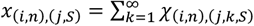.

For each age group 1 ≤ *i* ≤ *M*, let *S*_*i*_, *E*_*i*_, *I*_*i*_, *R*_*i*_ denote the fraction of the population that has age *i* and infection state *S, E, I*, or *R*. Note that *S*_*i*_ + *E*_*i*_ + *I*_*i*_ + *R*_*i*_ = 1. Note that a *sexually active individual* is of age *j* with probability

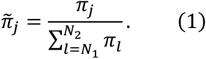

For convenience of notation, we let the vector **π**_**S**_ be

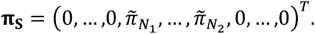

The size-biased degree distribution is

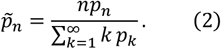

A key variable is

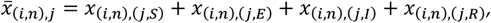

which is interpreted as the probability that a binding site is susceptible with a) age *i* b) degree *n*, and c) a partner of age *j*.

First, we consider the equations for the variables *x*_(*i,n*)(*j,y*)_ and 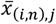. To derive the equations for these variables, we consider the events that can occur for susceptible binding sites:

1. A susceptible binding site with an infectious partner becomes infected via sexual contact at rate β. If that happens, the binding site “leaves” the *x*-system (meaning that the *x* variables form a defective probability distribution).
2. The infectious partner of the susceptible binding site can recover before infecting the binding site under consideration, this occurs at rate *γ*.
3. A susceptible binding site can have an exposed partner; this exposed partner becomes infectious at rate *σ*.
4. The susceptible individual to whom the binding site belongs can become infected through non-sexual contact, and this occurs at rate *pλ*_*i*_ with

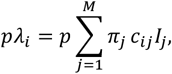

and affects each of the binding sites of the individual at the same time. Because of the independence of binding sites, the per-binding-site rate for an individual with degree *n* is *pλ*_*i*_/*n* = *p* ∑_*j*_ *π*_*j*_ *c*_*ij*_*I*_*j*_/*n*.
5. When a susceptible binding site is connected to a susceptible partner, the susceptible partner can get infected through either sexual contact with one of his/her other partners. The rate at which this occurs is explained below.
6. When a susceptible binding site is connected to a susceptible partner, the susceptible partner can get infected through non-sexual contact in the population. The rate at which this occurs is explained below.

Consider event (5). Transmission through sexual contact occurs at rate β per partner. Therefore, the rate at which event (5) occurs is β multiplied by the mean number of partners *Λ*_(*j,k*)_ of a susceptible partner *v*, where *v* has age *j* and degree *k*, and is a partner of a susceptible individual *u*. The fraction of individuals that a) are susceptible, b) are of age *j*, c) have degree *k*, and d) have a susceptible partner of age *i* and degree *n* as well as at least one infectious partner, is given by:

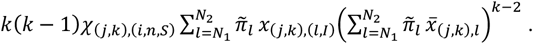

In contrast, the fraction of individuals who are susceptible, of age *j* and degree *k*, and who have a susceptible partner of age *i* and degree *n* is

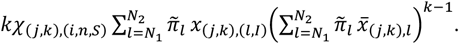

Note that these expressions use the independence of binding sites of a susceptible individual, allowing us to translate probabilities at the binding site level to probabilities at the individual level (see [16] for details). Therefore, the expected number of infectious partners of a susceptible partner *v* with degree *k* is

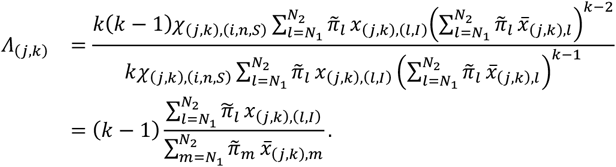

Finally, we determine the rate at which event (6) occurs. Again, suppose *v* has degree *k* then the probability that *v* is susceptible given *u* is 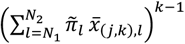. The rate at which *v* gets infected through non-sexual contacts is *λ*_*j*_. Therefore, the rate at which (6) occurs is

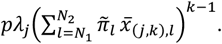

Putting the rates for the events (1)-(6) together yields the following (infinite-dimensional) system of equations for the *x* system for the sexually active ages *i, j* = *N*_1_, … *N*_2_:

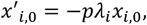

and for *n* ≥ 1,

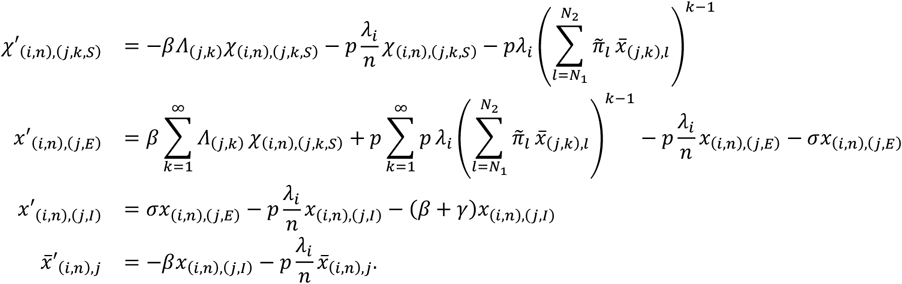

Next, for ages 1 ≤ *i* < *N*_1_ and *N*_2_ < *i* ≤ *M*, individuals are sexually inactive, so infection can only occur through non-sexual contacts:

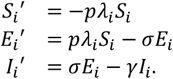

For the sexually active individuals, the equations for 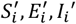 are derived as follows. An individual of age *N*_1_ ≤ *i* ≤ *N*_2_ that belongs to the sexually active group can be either of degree 0 or of degree *n* > 0. An individual of age *i* of degree *n* > 0 is susceptible with probability 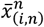, where 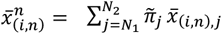. The probability that an individual is of degree *n* is *p*_*n*_. Therefore, 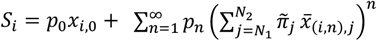. Differentiation with respect to time yields

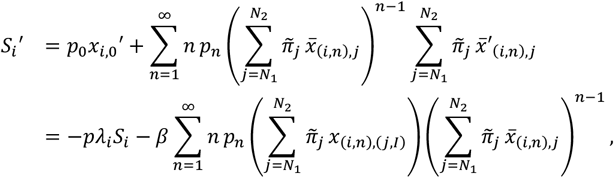

whereas the equations for *E*_*i*_′, *I*_*i*_′ are

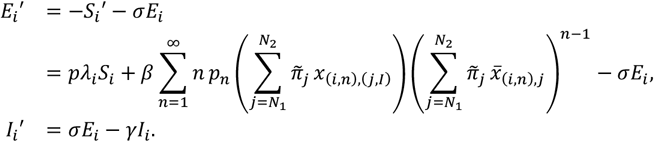

The ‘far past’ conditions are 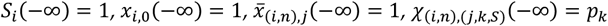, and all other variables are equal to zero.

##### 1.1 Next generation matrices

###### 1.1.1 Epidemiological interpretation

We derive the next generation matrix (NGM) directly from the epidemiological interpretation of how new cases can be caused by one newly infected case. We prove in Section 1.1.3 below that this derivation corresponds to linearization of the system of ODE. We characterize *R*_0_ as the dominant eigenvalue of an NGM ***K*** that contains both age and transmission route (transmission via sexual or non-sexual contact). The matrix ***K*** is therefore 2*M* × 2*M* dimensional. We characterize the elements of ***K*** as follows. Let *s* denote transmission via sexual contact, *ns* transmission via non-sexual contact. An element of the matrix ***K*** is denoted by *K*_(*j,y*),(*i,z*)_, with *i, j* = 1, …, *M* and *y, z* = *s, ns*.

First of all, some elements of ***K*** are zero. A newly infected individual in ages 1 ≤ *i* < *N*_1_, *N*_2_ < *i* ≤ *M* can only transmit via non-sexual contact *and* his/her infector could only have transmitted via non-sexual contact. Therefore *K*_(*j,ns*),(*i,s*)_ = 0 = *K*_(*j,s*),(*i,s*)_ = *K*_(*j,s*),(*i,ns*)_, 1 ≤ *i* < *N*_1_, *N*_2_ < *i* ≤ *M* and *j* = 1, …, *M*. Similarly *K*_(*j,s*),(*i,ns*)_ = 0 = *K*_(*j,s*),(*i,s*)_ for 1 ≤ *j* < *N*_1_, *N*_2_ < *j* ≤ *M, i* = *N*_1_ + 1, …, *N*_2_.

Now, there are the non-zero elements of ***K***. First consider a newly infected individual of age *i* who was infected via non-sexual contact. We are interested in the expected number *K*_(*j,ns*),(*i,ns*)_ of secondary cases of age *j* caused via non-sexual contacts by a newly infected individual of age *i* who was himself/herself infected via non-sexual contact. The expression follows the familiar form from mass action models:

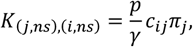

with *i, j* = 1, …, *M*. The expression is interpreted as follows: a newly infected individual is infectious for mean period 1/*γ*, contacts with age group *j* are made at rate *c*_*ij*_*π*_*j*_, and the probability of transmission upon contact with a susceptible individual is *p*.

Next, we consider elements *K*_(*j,ns*),(*i,s*)_. Note that non-sexual contacts are made indepenent of sexual contacts. Therefore, the expected number of secondary cases via non-sexual contact that are of age *j* is independent of the transmission route of the newly infected individual. Hence,

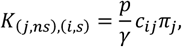

for *i* = *N*_1_, …, *N*_2_, *j* = 1, …, *M*.

Then consider elements *K*_(*j,s*),(*i,s*)_, *i, j* = *N*_1_, …, *N*_2_. Note that a newly infected individual who was infected via sexual contact has degree *n* with probability 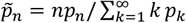, i.e., the size-biased degree distribution. The probability that a partner is of age *j* is 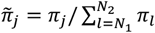. The probability that transmission to a partner occurs before the newly infected individual recovers is β/(β + *γ*). Therefore,

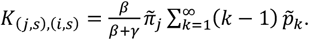

Finally, we consider the expected number of secondary cases *K*_(*j,s*),(*i,ns*)_ of age *j*, caused via sexual contact by a newly infected individual of age *i* who was him/herself infected via non-sexual contact:

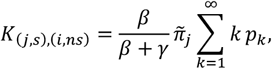

with *i, j* = *N*_1_, …, *N*_2_. This expression can be interpreted as follows: an individual of age *i* and degree *k* who is newly infected via non-sexual contact has *k* susceptible sexual partners, the probability that he/she transmits to a partner before recovery is 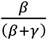, and the probability that the partner is of age *j* is 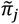.

Putting all these elements together, we can decompose the NGM ***K*** into four *M* × *M* matrices:

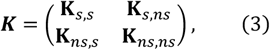

where the four submatrices are as follows. Let the vector **1**_**S**_ be the indicator function for sexually active age groups such that 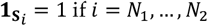 and 0 otherwise. Note that

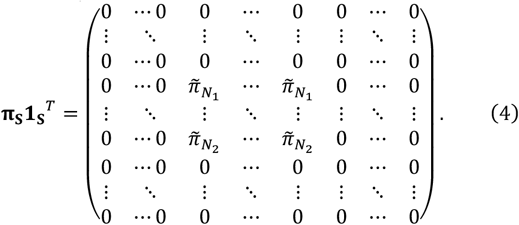

Then

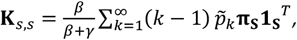

and

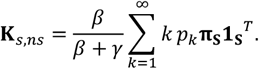

The submatrices involving secondary cases infected via the non-sexual route are

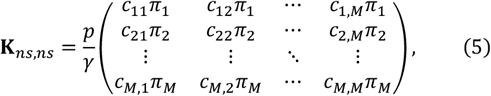

and

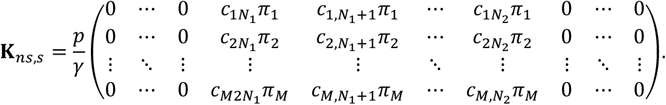

###### 1.1.2 Reductions of the NGM

As sexual partnerships are formed independent of age, we can reduce the NGM (3) by averaging over age in NGM elements involving transmission via sexual contact. This yields the NGM

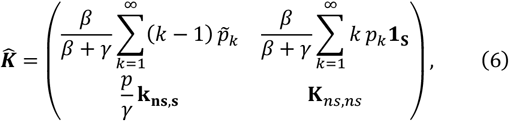

where

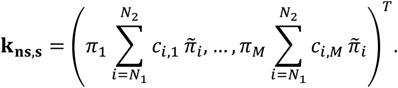

We provide the details for this reduction of *K* into 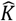 in Section 1.1.3.2.

As non-sexual contacts are age structured, i.e. the contact rates *c*_*ij*_ are dependent on *i* and *j*, we cannot further reduce the NGM. However, note that in the absence of age structure, the NGM 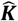 (6) can be reduced to

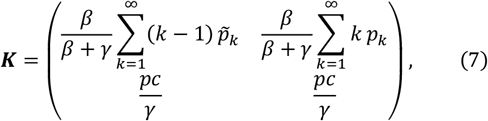

which corresponds to the NGM of [18] for the model without age structure. Note that the NGM (7) differs from the related model of [19], where a newly infected individual with *n* susceptible sexual partners generates an expected number of *β/γn* secondary cases via sexual contact rather than β/(β + *γ*)*n* as in our model. In the absence of both age structure and non-sexual contacts, i.e. in a model with only network structure, one obtains a one-dimensional NGM equal to the basic reproduction number 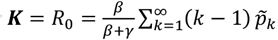, e.g. [17]. Finally, in the absence of the network structure, the NGM reduces to **K**_*ns,ns*_ of (9), which corresponds to the NGM of an age-structured mass action SEIR model [20].

###### 1.1.3 R_0_ is a threshold parameter for the model

The NGM ***K*** is derived directly from the epidemiological interpretation given in Section 1.1.1. We show that the dominant eigenvalue *R*_0_ of ***K*** is indeed a threshold parameter for the model in the sense that it determines the stability of the disease free steady state. To demonstrate this, we linearize the system around the disease free steady state, construct the NGM 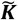 from the linearized system, and show that both 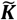 and ***K*** can be reduced to the same NGM with small domain 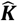 given by (6). This proves that all three matrices 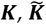 and 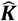 have the same dominant eigenvalue *R*_0_. Since the dominant eigenvalue of 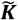 is a threshold parameter for the disease free steady state of the model, *R*_0_ is indeed the threshold parameter that we are after.

###### 1.1.3.1 Linearization of the ODE system

We prove that *R*_0_ is a threshold parameter of the system by linearization around the disease-free steady state, following the approach in [17]. In this linearization, one finds a decoupled system for the variables *E*_*i*_, *I*_*i*_, *x*_(*i,n*),(*j,E*)_, *x*_(*i,n*),(*j,I*)_, which can be further reduced to a linearized system for the variables 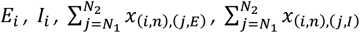. Additionally, in the linearization the variables 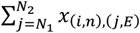 and 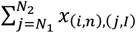 are independent of *n*, which allows further reduction by averaging over *n*. We then split the linearized system into a transmission matrix ***T*** and a transition matrix ***Σ***, yielding the NGM with large domain 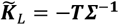. The matrix 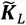 can be reduced to 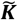 by considering states-at-infectiousness. This yields a 2*M* × 2*M* matrix. One can interpret the NGM 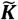 in terms of “reproduction opportunities” (see [17], section 4.3).

We characterize *R*_0_ as the dominant eigenvalue of an 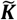. The 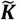 contains both age and transmission route (transmission via sexual or non-sexual contact), and is therefore 2*M* × 2*M* dimensional. We characterize the elements of 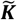 as follows: let *s* denote transmission via sexual contact, *ns* transmission via non-sexual contact, elements 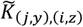 of the matrix 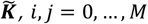 and *y, z* = *s, ns*. A newly infected individual in age group 1 ≤ *i* < *N*_1_, *N*_2_ < *i* ≤ *M* can only transmit via non-sexual contact *and* his/her infector must also have transmitted via non-sexual contact. Therefore 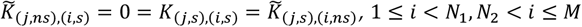 and *j* = 1, …, *M*. Similarly, 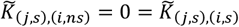 for 1 ≤ *j* < *N*_1_, *N*_2_ < *j* ≤ *M, i* = *N*_1_, …, *N*_2_. Linearization yields the following NGM elements:

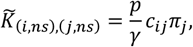

with *i, j* = 1, …, *M*,

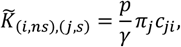

for *i* = *N*_1_, …, *N*_2_, *j* = 1, …, *M*,

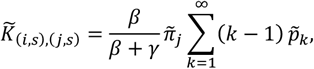

for *i, j* = *N*_1_, … *N*_2_,

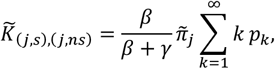

for *j* = *N*_1_, …, *N*_2_.

Putting all these elements together, we can decompose the NGM ***K*** into four *M* × *M* submatrices:

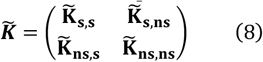

where the four matrices are as follows.

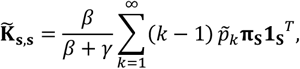

and

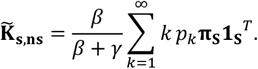

The submatrices involving secondary cases infected via non-sexual contact are

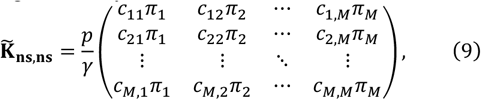

and

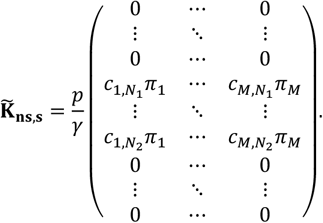

###### 1.1.3.2 Reduction to 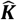

We provide the calculations for the reduction of ***K*** into 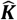, where ***K*** and 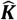 are given by (3) and (6). The NGM 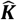 can be interpreted as the NGM with small domain of ***K*** (e.g. [21]). Here the reduction is in the sense that ***K*** and 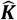 have the same dominant eigenvalue.

Let ***C*** be the following (1 + *M*) × (*M* + *M*) matrix:

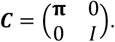

Let ***R*** be the following (*M* + *M*) × (1 + *M*) matrix:

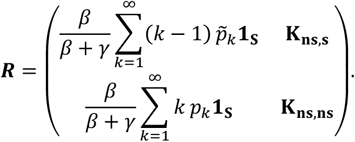

Then ***RC*** = ***K***^*T*^ and 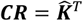.

Similarly, we can reduce the NGM 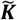 to 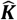. Let 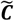 be the following (1 + *M*) × (*M* + *M*) matrix:

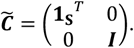

Let 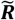 be the following (*M* + *M*) × (1 + *M*) matrix:

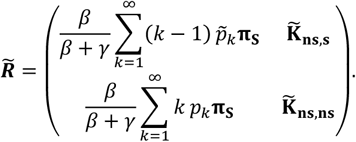

Then 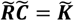 and 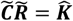.

###### 1.1.3.3 Dominant eigenvalue

The system of ODEs describing the model of the sexual network together with non-sexual contacts can be linearized around the disease free steady state. The matrix for the linearization can be decomposed into a transmission matrix ***T*** and a transition matrix ***Σ*** such that − 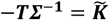. Since both 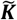 and ***K*** have 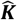 as the NGM with small domain, the dominant eigenvalue *R*_0_ of ***K*** is a threshold parameter with threshold value of one for the stability of the disease free steady state of our model. Furthermore, *R*_0_ has the usual interpretation as the expected number of secondary cases caused by one typical newly infected individual.

#### 2 Newly infected individuals over time

##### 2.1 Number of sexual partners of newly infected individuals over time

Individuals of age group *i* become infected via non-sexual contact at rate *pλ*_*i*_*S*_*i*_ and via sexual contact at rate 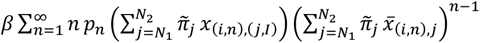 (see ODE for *E*_*i*_).

The rate at which an individual of age *i* and degree *n* gets infected via non-sexual contact is *p*_*n*_*pλ*_*i*_*S*_*i*_ since the route of transmission is independent of the degree. The probability that a newly infected individual of age *i*, infected via non-sexual contact, has degree *n* is

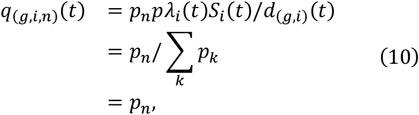

where *d*_(*g,i*)_(*t*) : = ∑_*n*_ *p*_*n*_ *pλ*_*i*_(*t*)*S*_*i*_(*t*) = *pλ*_*i*_(*t*)*S*_*i*_(*t*) is the (time-varying) normalizing constant.

An individual of age *i* and of degree *n* gets infected via sexual contact at rate 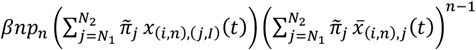. Therefore, the probability that a newly infected individual of age *i* who was infected via sexual contact, has degree *n* is given by

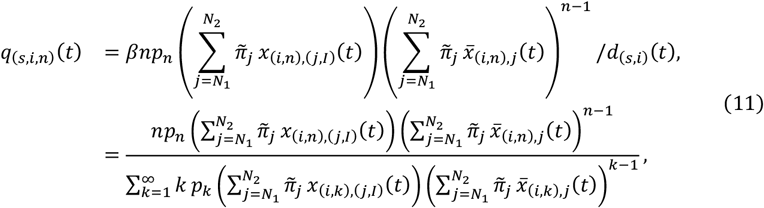

where 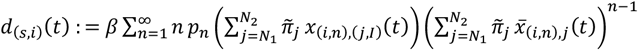 is the normalizing constant.

##### 2.2 Time-varying NGM

We derive the time-varying NGM from the interpretation similar to Section 1.1.1. With a little abuse of notation, we again use ***K*** to denote the time-varying NGM, but we now explicitly include the time dependence: ***K***(*t*).

First, consider a newly infected individual of age *i* who was infected via non-sexual contact. We are interested in the expected number *K*_(*j,ns*),(*i,ns*)_(*t*). We now need to take into account the fraction of susceptible individuals of age *j* at time *t*:

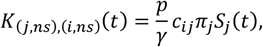

with *i, j* = 1, …, *M*. Similarly,

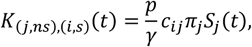

for *i* = *N*_1_, …, *N*_2_, *j* = 1, …, *M*.

Next, we consider a newly infected individual of age *i*, infected via sexual contact, and derive the expected number of secondary cases *K*_(*j,s*),(*i,s*)_(*t*), *i, j* = *N*_1_, …, *N*_2_. At time *t*, a newly infected individual who was infected via sexual contact has degree *n* with probability *q*_(*s,i,n*)_(*t*) (see (11)). At the moment of infection, partners are independent of one another. We can consider the probability that a partner of age *j* of a newly infected individual has degree *k*. This probability is given by the size-biased degree distribution 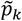. He/she is susceptible at time *t* with probability 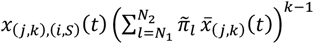 (one of his/her *k* partners was recently susceptible and has age *i*, the other *k* − 1 binding sites are susceptible). Therefore, the probability that a partner of age *j* of a newly infected individual of age *i* at time *t* is susceptible is equal to

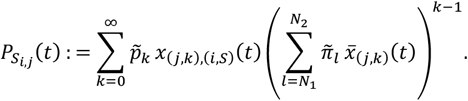

Note that 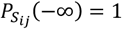, corresponding to all partners being susceptible in the disease-free steady state. The time-dependent NGM element *K*_(*j,s*),(*i,s*)_(*t*) is:

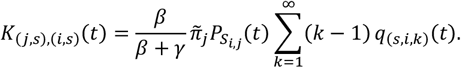

Finally, the expected number of secondary cases *K*_(*j,s*),(*i,ns*)_(*t*) is:

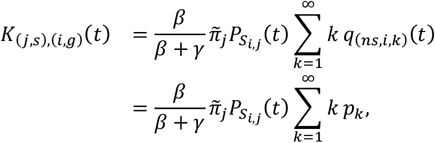

with *i, j* = *N*_1_, …, *N*_2_, and *q*_(*ns,i,k*)_ is given by (10).

As in Section 1.1.1, all other elements are equal to zero. Putting everything together in a matrix of the form (3) we obtain the time-varying NGM ***K***(*t*).

##### 2.3 Mean degree of a newly infected individual over time

To obtain the mean number of partners of a newly infected individual, we proceed as follows. Averaging over age yields the rate at which an individual of degree *n* gets infected:

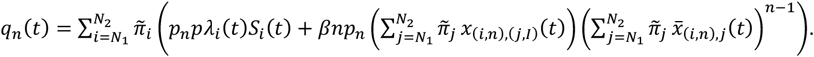

By normalizing this distribution (*q*_*n*_(*t*))_*n*_ and averaging over *n*, we obtain the mean degree of a newly infected individual at time *t*.

#### 3 Perturbation analysis of NGM

##### 3.1 Framework

In the following sections, we quantify the expected impact of targeted interventions on the time-varying NGM **K**(*t*) (Section 2.2) and consequently on its dominant eigenvalue *R*(*t*). From hereon, we drop explicit writing of *t* and use **K** and *R* for notational simplicity. The general principle is to identify the group that yields the largest expected reduction in *R* when a single unit of vaccines is allocated to that group. We begin by introducing the sensitivity and elasticity of matrices and their epidemiological interpretations. We then formulate the expected change in the NGM **K** for different targeted vaccination strategies. This approach has been widely used in demography and ecology [22], and as a tool to analyze the NGM in epidemiology [21], [23]−[25]. Here, we focus on vaccine allocation to illustrate this framework, although it can be extended to other interventions such as school or workplace closures.

##### 3.2 Sensitivity and elasticity of NGM

Targeted interventions influence the entries of the NGM **K**. We denote the matrix entries of **K** by *k*_*mn*_, with *m, n* = 1, …, 2*M*, representing all combinations of age (across *M* age groups) and transmission route (sexual *s* or non-sexual *ns*). The sensitivity of *R* with respect to a change in entry *k*_*mn*_ is expressed as

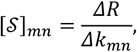

and the elasticity is given by

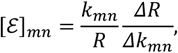

or in matrix form

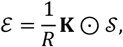

where ⊙ represents the Hadamard (element-wise) product. The cumulative elasticity for the column *ℓ* is obtained by summing over the elasticities [ℰ]_*mℓ*_ across all groups *m*:

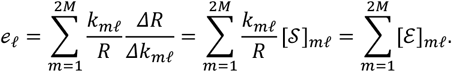

The cumulative elasticity *e*_*ℓ*_ measures the proportional contribution of column *ℓ* to the change in *R* given the same perturbation to every entry *k*_*mn*_. If we apply it to an age-stratified NGM with a single transmission route, the cumulative elasticity can be interpreted as the total contribution from a specific age group to the overall transmission potential *R*.

In our setting, an NGM element represents the age- and transmission route-specific number of secondary infections. We define the age-specific contribution to *R* by summing the cumulative elasticities of age group *i* for two transmission routes:

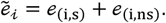

Similarly, we define the cumulative sensitivity of age group *i* as 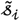.

##### 3.3 Targeted vaccination

###### 3.3.1 Number of vaccines, vaccination coverage, and effective vaccination coverage

Let *u*_*i,n*_ denote the number of vaccines (i.e., the number of individuals who complete the full vaccine series) allocated to the group of age *i* with degree *n* (hereafter called the group (*i, n*)). The total number of vaccines allocated to the age group *i* is given by:

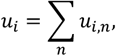

and the total number of vaccines allocated across the entire population is

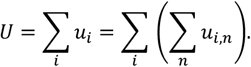

We define the vaccination coverage for group (*i, n*) as the proportion of vaccinated individuals:

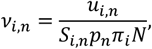

where *S*_*i,n*_ is the proportion susceptible in group (*i, n*) and *N* is the total population. By considering vaccine efficacy *ε*_*i,n*_ (VE) against infection in group (*i, n*), *ε*_*i,n*_the proportion of effectively vaccinated individuals (i.e., effective vaccination coverage) in group (*i, n*) is given by *ε*_*i,n*_*ν*_*i,n*_. In the present study, we assume a uniform VE for all age groups: *ε*_*i,n*_ = *ε*.

###### 3.3.2 Impact of targeted vaccination strategies

Here, we consider the unit change in the total number *ΔU* of vaccines allocated. We compute the unit change *Δν*_*i,n*_ in the vaccination coverage in group (*i, n*), if the single unit of vaccines is allocated to that group.

If the vaccination is at random, vaccines will be allocated proportional to the relative population size of the age group *i*:

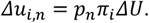

The unit change in the vaccination coverage is given by

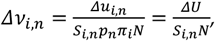

where *p*_*n*_ is the proportion with degree *n, π*_*i*_ is the proportion of age *i, S*_*i,n*_ is the proportion of suscepibles in age group *i* with degree *n*, and *N* is the total population size. This represents the situation where all groups (*i, n*) receive the same number of vaccines. The unit change in the vaccination coverage *Δν*_*i,n*_ is further used to derive an NGM under vaccination (see section 3.4 below).

##### 3.4 NGM with vaccination

###### 3.4.1 Change in the proportion of susceptibles in group (i, n)

Using the unit change in the effective vaccination coverage *εΔν*_*i,n*_, the unit change in the proportion of susceptibles in group (*i, n*) is expressed as:

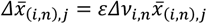

and this holds for any partner of age group *j*. In the next section, we relate this unit change to the change in elements of the NGM and quantify the cumulative sensitivity and elasticity.

###### 3.4.2 Change in the elements of the NGM

Let 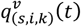 denote the probability that a newly infected individual of age *i* infected through sexual contact (*s*) has degree *n* under vaccination (*v*). First, if age group *i* is targeted for vaccination, the probability that a binding site is susceptible with age *i* and degree *n* and has a partner of age *j* after a unit increase in effective vaccination coverage is given by:

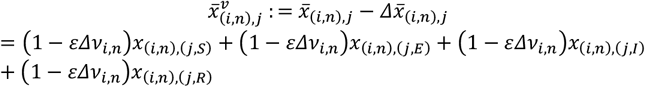

because the vaccine reduces the proportion of susceptibles in age *i*, independent from the partner’s infection status (i.e., the same reasoning for computing 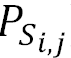).

Second, this change leads to the change of 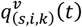, and it is expressed as:

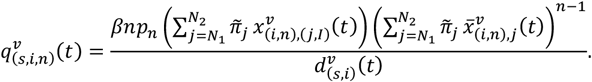

By substituting (1 − *εΔν*_*i,n*_)*x*_(*i,n*),(*j,I*)_ and 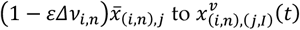 and 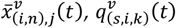 is rearranged as

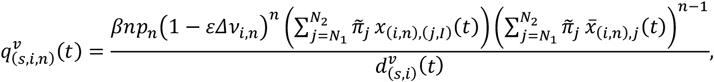

and 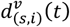 is rearranged as

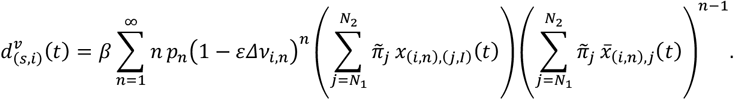

For age groups that are not targeted, the probabilities *q*_(*s,i,n*)_(*t*) remain the same.

Next, we consider 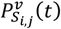, the probability that a partner of age *j* of an individual in age group *i* is susceptible after vaccination. If age group *j* is not targeted, the probability remains unchanged, i.e.,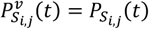. If age group *j* is targeted, then:

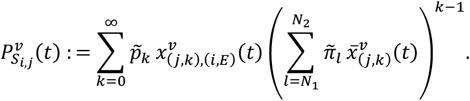

By using 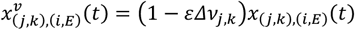 and 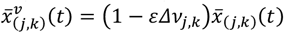, we obtain

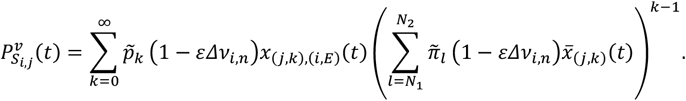

Lastly, for transmission via non-sexual contact, only the proportion of susceptibles in age group *i* is affected. The updated proportion of susceptibles after vaccination is:

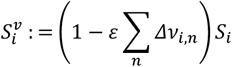

Using the result of section 2.2, the changes in susceptible-related terms (i.e., 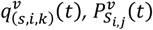 and 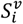) directly propagate into the updated NGM after vaccination, **K**^*v*^, which incorporates the effect of vaccination on transmission potential. From this, one can compute the post-vaccination sensitivities and elasticities of the system, analogous to the framework described in Section 3.2.

